# Identification of collagen features predictive of recurrence following radiotherapy for localised prostate cancer: a retrospective case control analysis

**DOI:** 10.64898/2026.07.16.26358234

**Authors:** Robert P. Jenkins, Xiao Fu, Sara Waise, Monisha Dewan, Clare Griffin, Christine Stuttle, Clare Cruickshank, David Dearnaley, Isabel Syndikus, Emma Hall, Erik Sahai, Anna Wilkins

**Author notes:** Joint senior and corresponding authors Corresponding authors contacts: Anna Wilkins, The Institute of Cancer Research, 123 Old Brompton Road, London SW7 3RP Erik Sahai, The Francis Crick Institute, 1 Midland Road, London NW1 1AT.

## Abstract

**Background:** Changes in the extracellular matrix (ECM) are a recognised feature of aggressive prostate cancer, but they are not exploited in clinical decision-making. We aimed to develop automated quantitative ECM parameters to facilitate risk stratification for localised prostate cancer.

**Methods:** 378 quantitative ECM parameters were derived from picrosirius red-stained diagnostic prostate biopsies in a cohort of 422 patients, matched 1:1 for recurrence, recruited to the CHHiP (Conventional or Hypofractionated High Dose Intensity Modulated Radiotherapy in Prostate Cancer) trial of radiotherapy fractionation for localised prostate cancer. These ECM parameters comprehensively described fibre architecture, gaps and ECM texture. Machine learning models at the level of both individual image tiles and patients defined how ECM parameters related to tumour versus normal prostate, Gleason grade group and recurrence. Shapley analysis was used to interpret ECM feature importance and develop signatures associated with recurrence.

**Results:** Specific ECM patterns identified tumour versus normal prostate, Gleason pattern 4 versus 3 and recurrence. ECM patterns associated with recurrence were enriched in Gleason 4+3 patients, versus Gleason 3+4 patients. Shapley analysis revealed that biopsies from patients with recurrence had smaller more elongated gaps between fibres, with finer grained ECM texture and lower ECM homogeneity than less recurrent regions.

**Interpretation:** Quantitative automated analysis of ECM architecture can inform probability of prostate cancer recurrence after radiotherapy; Features relating to ECM gap size and texture are of particular relevance.

**Funding:** This work was funded by Prostate Cancer Research, Cancer Research UK and the Francis Crick Institute.

## Background

Worldwide, over 1.46 million men were diagnosed with prostate cancer in 2022, with approximately 400,000 deaths occurring from the disease in the same year (1). Current clinical challenges in the management of prostate cancer include the need for improved treatment stratification in localised disease – here, a substantial proportion of men with indolent disease do not need definitive treatment whereas other men with aggressive tumours need treatment intensification. In more advanced disease, new therapeutic strategies are urgently needed to improve survival.

The role of non-cancerous cells within the tumour microenvironment (TME) in shaping prostate cancer evolution is increasingly recognised (2–4). Cancer-associated fibroblasts (CAFs) and the associated extracellular matrix (ECM), that is predominantly produced by such fibroblasts, are important TME constituents. Feedback between fibroblasts and the matrix creates diverse higher order matrix patterns with important impacts on tumour behaviour (5). Emerging data indicate that CAFs and the ECM can drive aggressive tumour behaviour across prostate cancer stages (2–4, 6, 7). For example, in localised disease, pathologists have provided detailed histological descriptions of a “stromogenic” or “reactive” prostate cancer variant characterised by excessive deposition of ECM and hyperproliferation of myofibroblasts between angular deformed tumour glands (8). This variant is associated with increased recurrence across international cohorts (9, 10). Furthermore, in pre-clinical prostate models and human samples, the geometry of ECM fibres is related to metastatic frequency (11). Here, a more aligned matrix is seen in tumours with higher metastatic potential than the non-aligned matrix seen in tumours that do not metastasize.

Recent mechanistic insights relevant to these observations include the identification of CAF to tumour cell crosstalk via neuregulin 1 (NRG1) and HER3, which promotes anti-androgen resistance (12). Elsewhere, TGFβ has been shown to induce stromal fibroblast reprogramming to an SPP1+ CAF phenotype upon anti-androgen treatment; subsequent SPP1-ERK paracrine signalling can promote castration resistance in tumour cells (2). Histological and transcriptomic analysis in patient tumours indicates that both of these mechanisms are clinically relevant and contribute to poor outcomes across different stages of prostate cancer, including castration-resistant disease (2, 12).

Despite the above insights, very limited quantitative evaluation of stromogenic patterns has restricted clinical application. The detailed histopathological descriptions of stromogenic tumours outlined above are time consuming and subjective. Transcriptomic signatures of reactive prostate cancer can identify aggressive tumours but are not currently feasible or affordable to apply in the clinic (4). A number of machine learning methods show promise in predicting histological features, such as Gleason grade group, as well as identifying recurrence, in prostate cancer (13–17). These have mainly been developed using Haematoxylin and Eosin (H&E) stained diagnostic tissue and show very good ability to identify tumour regions and assign Gleason grade group. However, they have tended to focus on tumour glands and exclude the stroma, in part because of the limited biological variation detected in Eosin-stained stroma, in contrast to Haematoxylin-stained tumour glands.

Outside of prostate cancer, new methods for quantitative evaluation of the ECM have been identified, including TWOMBLI and CT-FIRE, amongst others (18, 19). To our knowledge, these have not been applied to well-sized human prostate cancer cohorts and it is likely that we currently underestimate the complexity of matrix patterns in prostate cancer and their influence on disease phenotype.

## Results

### Definition of clinical cohort with balanced recurrence status and Gleason grade group

To explore whether patterns of collagen organisation can predict the likelihood of prostate cancer recurrence, we established a pipeline for acquiring and analysing collagen I images and applied it to patients with localised prostate cancer (Figure 1). Patients were selected from the randomised phase 3 CHHiP trial of radiotherapy fractionation (ISRCTN97182923 - summarised Figure 1A & Figure S1A). In this study, tumours were predominantly of National Comprehensive Cancer Network (NCCN) intermediate risk group (20), with a low recurrence rate of 20-24% at 10 years follow up (21). In light of this, a nested case control methodology was used to define the study cohort. 211 cases with recurrence, and 211 controls without recurrence were matched based on time to biochemical or clinical recurrence and centrally- assigned Gleason score (22). This matching minimised differences in Gleason grade group confounding ECM parameters. Histology review of additional sections taken from tissue blocks led to the exclusion of 86 cases with no tumour present and 19 cases with intraductal carcinoma and no obvious invasive cancer, thus sections from 317 patients were taken forward for staining with Picrosirius Red and Haematoxylin (PSR+H), which enables visualisation of both stromal architecture and nuclear morphology. Of these, 4 had distorted morphology and were excluded. Following removal of patients that were classified as both control and then later a case, the final study cohort of 141 cases and 155 controls is shown in Figure 1B, alongside Gleason score categories, which were well balanced between cases (with recurrence) and controls (without recurrence).

**Figure 1.**
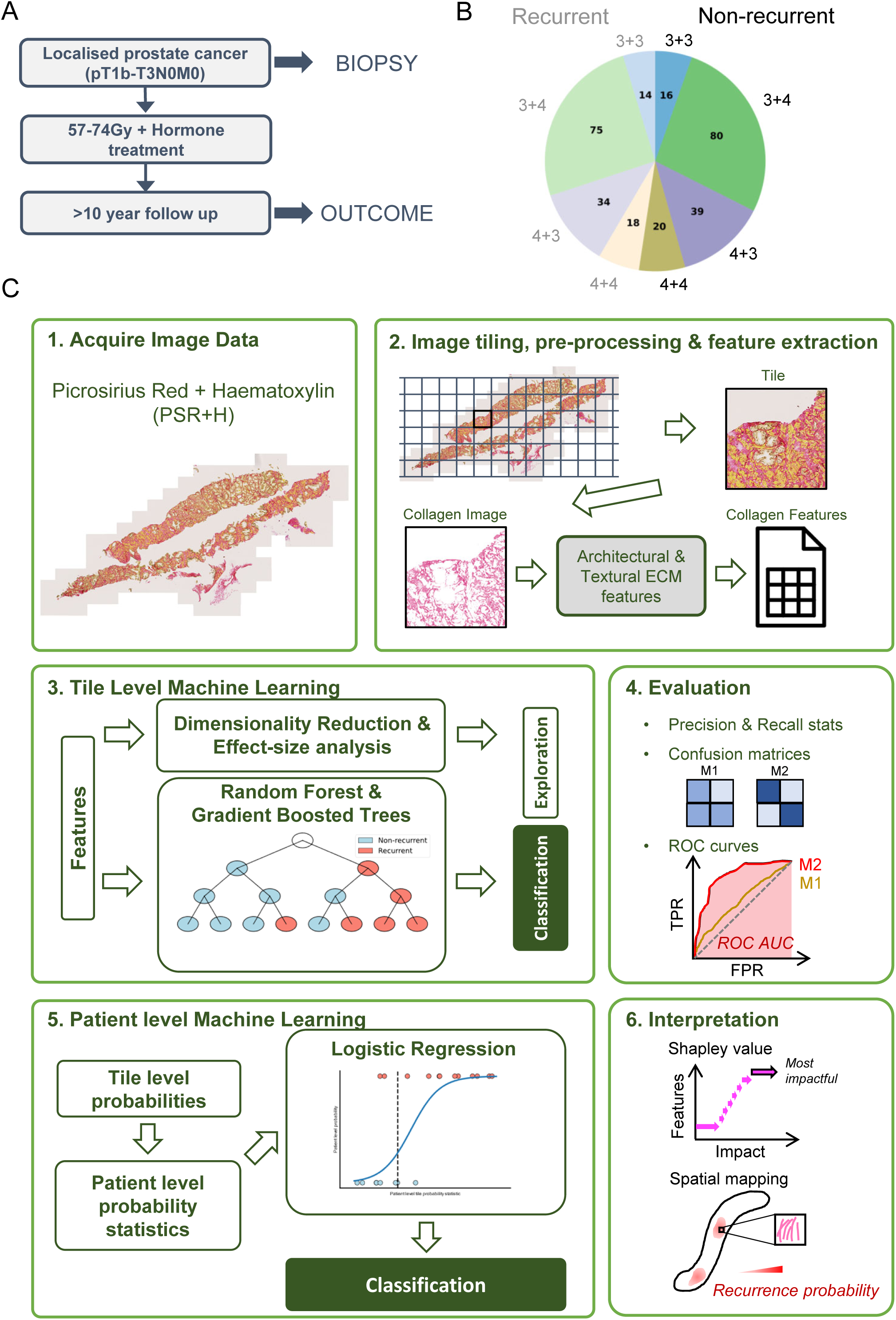
Schematics of clinical trial and machine learning pipelines. A: Schematic of biopsy and follow-up pipeline of CHHiP trial. B: Chart showing matched distribution of recurrence and Gleason grades at patient level. C: Schematic illustration of the spatial data analysis pipeline based on Picrosirius Red and Haematoxylin (PSR+H) stained images of prostate biopsies (1). Main components include (2) image pre-processing (e.g., tiling and colour deconvolution) and feature extraction across multiple data domains, (3) machine learning for exploration and classification of recurrence status at tile level, (4) model performance evaluation at tile and patient level, (5) input of tile level information into patient level machine learning models and (6) interpretation of most impactful features driving model classification of recurrence status.

### Image pre-processing and generation of multi-modal quantitative spatial features based on ECM organisation

A schematic illustration of the spatial data analysis pipeline based on picrosirius red and haematoxylin (PSRH) stained images of prostate biopsies is shown in Figure 1C. Following acquisition of PSRH-stained images, tissue masks and clinical tumour annotations were applied. This was followed by the overlay of a grid and extraction of 1000 pixel (220µm) square tiles from whole slide images (Figure 2A). Ultimately, this generated >40,000 tiles from the 296 patients. Tissue and tumour proportions varied widely between tiles as shown in Figures S2A-C, but the number of tiles per patient did not differ by recurrence status (Figure S2D). Figure 2B&C show how a tissue mask was applied and colour deconvolution to extract the PSR-stained collagen were performed, followed by image analysis to derive quantitative spatial features. These features directly related to discrete ECM fibres extracted via CT-FIRE (19), gaps between fibres (23), and fibre texture – the latter predominantly via Gray-Level Co-occurrence Matrix (GLCM) texture analysis. Figure 2D shows the full range of quantitative spatial features generated. Not all tiles were entirely filled with tissue; therefore, we explored whether this might confound our metrics or make them unreliable. Analysis of the variability in feature value relative to the proportion of tissue in a tile revealed that measurements were noisy in tiles with less than 25% tissue (Figure S2E). Following this analysis, we adopted a highly conservative approach and chose to restrict our analysis to tiles with >70% tissue. To remove the effects of varying tissue proportion in the remaining tiles, feature values were normalised according to a power-law fitted to the relationship between the metric and tissue proportion (Figure S2F). To maximise the biological robustness of extracted ECM features, we removed or combined features that were highly correlated (>0.999). This left a total of 378 ECM features from an initial set of 478 features.

**Figure 2:**
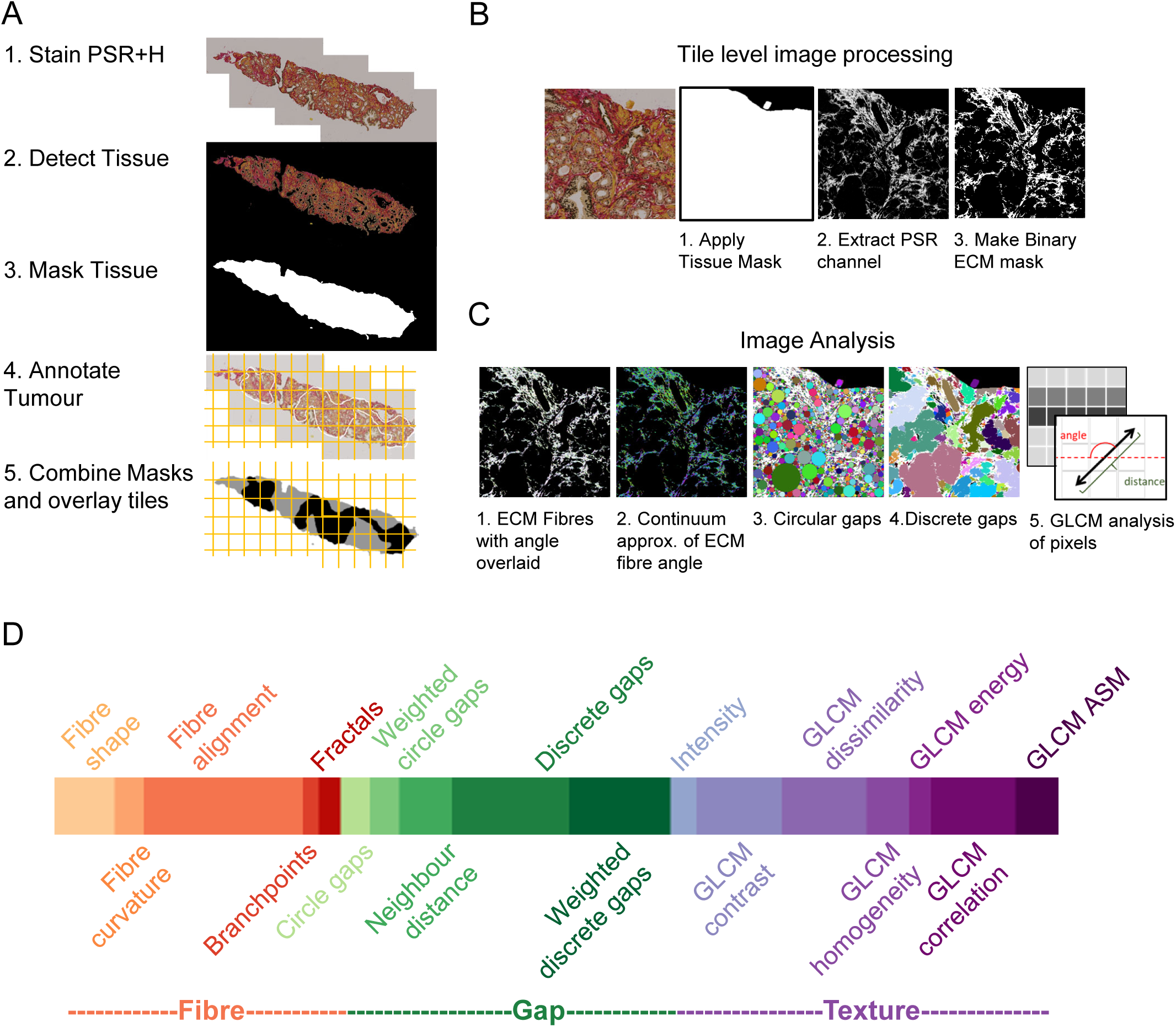
Distribution of data, pre-processing steps and feature generation. A: Schematic illustrating whole slide image (WSI) pre-processing steps – tissue detection using k-means clustering and tumour detection via clinical annotation. B: WSI are too large to analyse and so are tiled at various sizes. We used a tile size of 220 microns squared throughout. Once tiled, tiles are deconvolved to three channels. The PSR channel is used for analysis of extracellular matrix (ECM) alongside the tissue mask tile and binary ECM mask generated from the PSR channel. C: Analysis pipelines applied to deconvolved PSR channel – 1&2) Analysis of discrete ECM fibres following extraction via CT-FIRE, 3&4) analysis of inverse problem –the gaps between matrix and 5) analysis of texture using gray level co-occurrence matrices (GLCM). D: Final distribution of features used for analysis following feature robustness pipelines in Figure S2 and Methods.

### Discrimination of tumour, Gleason grade, and recurrence using ECM metrics

Dimensionality reduction of ECM features using UMAP is shown in Figure 3A, with the values of some exemplar metrics shown by colour variation in Fig. S3A. Exemplar tiles at various points in UMAP space illustrate normal tiles (blue box) and tumour tiles (black and grey boxes) with black boxes representing non-recurrent tumour tiles and grey boxes recurrent tumour tiles. Interestingly, there is no overall separation between normal or tumour regions or between tiles from recurrent or non-recurrent patients; although, tumour tiles frequently had higher UMAP1 and UMAP2 values (Figure 3A&B). Fig. S3B also highlights the inter-patient heterogeneity in both normal and cancerous prostate tissue. To determine which features best distinguished between normal and tumour, we compared the distribution for each feature between normal prostate and tumour tiles via a multiple testing corrected t-test and Cohen’s-d effect-size (Figure 3B). Many of the features were highly significant in terms of difference in mean, driven by the large numbers of tiles in each sample. However, the effect-size demonstrated that no individual feature could cleanly separate the distributions of the two origins (Figure 3B). The results demonstrate that even for the reasonably trivial task of separating tumour from normal regions, multiple features will most likely require consideration in unison.

**Figure 3:**
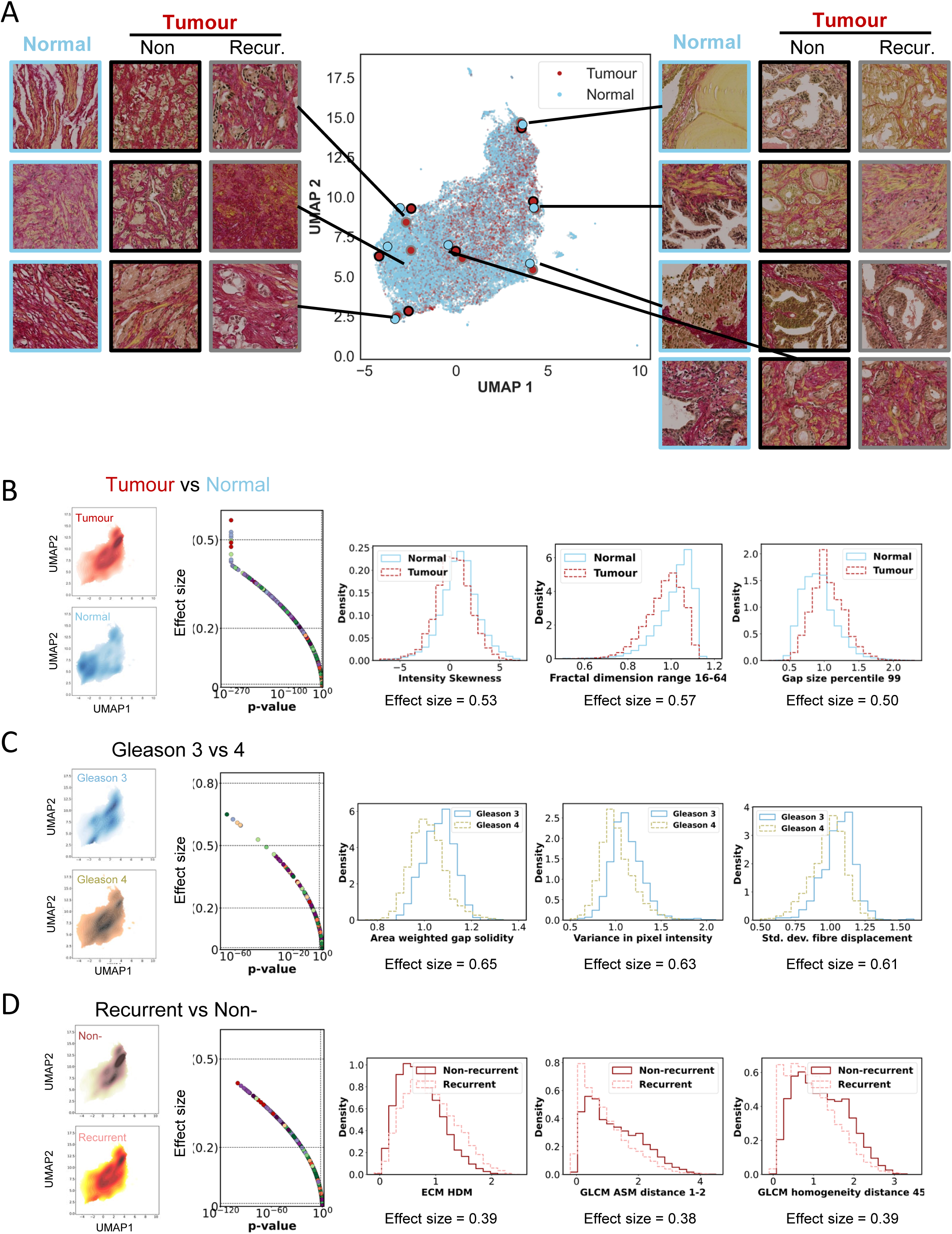
Tile level data exploration. A: UMAP dimensionality reduction with 100% tumour (red) versus 0% tumour tiles (blue) with exemplar tiles overlaid. Recurrent tumour tiles have a grey boundary and non-recurrent tumour tiles a black boundary. B: Left - Kernel Density Estimates (KDEs) of tumour tiles and normal tiles. Middle & right - Plot of False Discovery Rate (FDR) corrected p-value versus effect-size for all features of normal versus tumour and histograms of three features with largest effect-size. C: Left - KDEs of Gleason 3 and Gleason 4 tiles. Middle & right - Plot of FDR corrected p-value versus effect-size for all features of Gleason 3 versus Gleason 4 and histograms of three features with largest effect-size. D: Left - KDEs of recurrent and non-recurrent tiles. Middle & right - Plot of FDR corrected p-value versus effect-size for all features of recurrent versus non-recurrent and histograms of three features with largest effect-size.

We also used UMAP to explore how ECM pattern related to Gleason grade group 3 versus Gleason grade group 4, and observed a fairly wide spread of ECM features within individual Gleason categories (Figure 3C). This suggests that considerable biological variation in ECM features arises independently of Gleason grade group. Many features had highly statistically significant differences between Gleason grade. Interestingly, although there were fewer features showing moderate effect-size compared to tumour versus normal, a small subset reached moderate to large effect-size (highlighted in the histograms shown in Figure 3C).

Next, we explored how ECM features related to recurrence status. We observed that recurrence versus non-recurrence was well mixed in UMAP space (Figure 3A and S3B). Encouragingly, many ECM features were significantly different for recurrence versus non-recurrence with many demonstrating small to moderate effect-size (Figure 3D). Having said this, the smaller magnitude of differences between recurrent and non-recurrent tiles compared to tumour versus normal or Gleason 3 versus 4, indicates that predicting recurrence versus non-recurrence will be a harder problem to solve. We additionally considered if there might be differences in ECM in non-cancerous tissue in patients who recurred compared to those who did not (Fig. S3C). Figure S3C Shows that only very small differences were observed, arguing against a prostate-wide field effect being linked to recurrence. PTEN and Ki67 have been linked to recurrence after prostate radiotherapy (24); however, neither showed a clear association with ECM features (Figures S3D&E).

### Machine learning-based prediction of tumour and Gleason grade from matrix features

Next, we sought to train machine learning models to predict whether tiles were ‘normal’ (<10% tumour) or tumour (>90% tumour). Gradient boosted trees and random forests were compared using a nested cross validation approach with five inner and five outer folds (Figure S4A). The outer folds partitioned the data into five non-overlapping sets each containing 20% of the data. The process of model fitting was then carried out five times with each fold constituting an independent test set and the remaining 80% of data used for training each time. The inner folds split the corresponding training data for each outer fold further into five sets of training and validation data at 80-20 split. The inner folds were used to hyperparameter tune the models, generating five inner validation performance scores per hyperparameter combination. For a given outer fold, the hyperparameter combination with the highest mean score from inner folds was then selected, fitted to all training data for the outer fold and tested on the outer fold test set. Ultimately, the process generates five final models, one per outer fold, and consistent performance across folds indicates stable generalisability of the modelling approach. During stratification, all tiles from a single patient were within the same inner or outer fold, and as far as possible, the same proportion of recurrent and non-recurrent patients at each Gleason grade were placed in each fold. Stratification also accounted for number of tiles per patient, avoiding some folds being populated by far larger numbers of tiles than others. Performance was measured using ROC-AUC (Receiver Operating Characteristic Area Under the Curve). Figure 4A shows ROC curves from the gradient boosted tree to predict tumour versus normal prostate with a mean AUC of 0.87, while Figure S4B shows the slightly poorer random forest performance.

**Figure 4:**
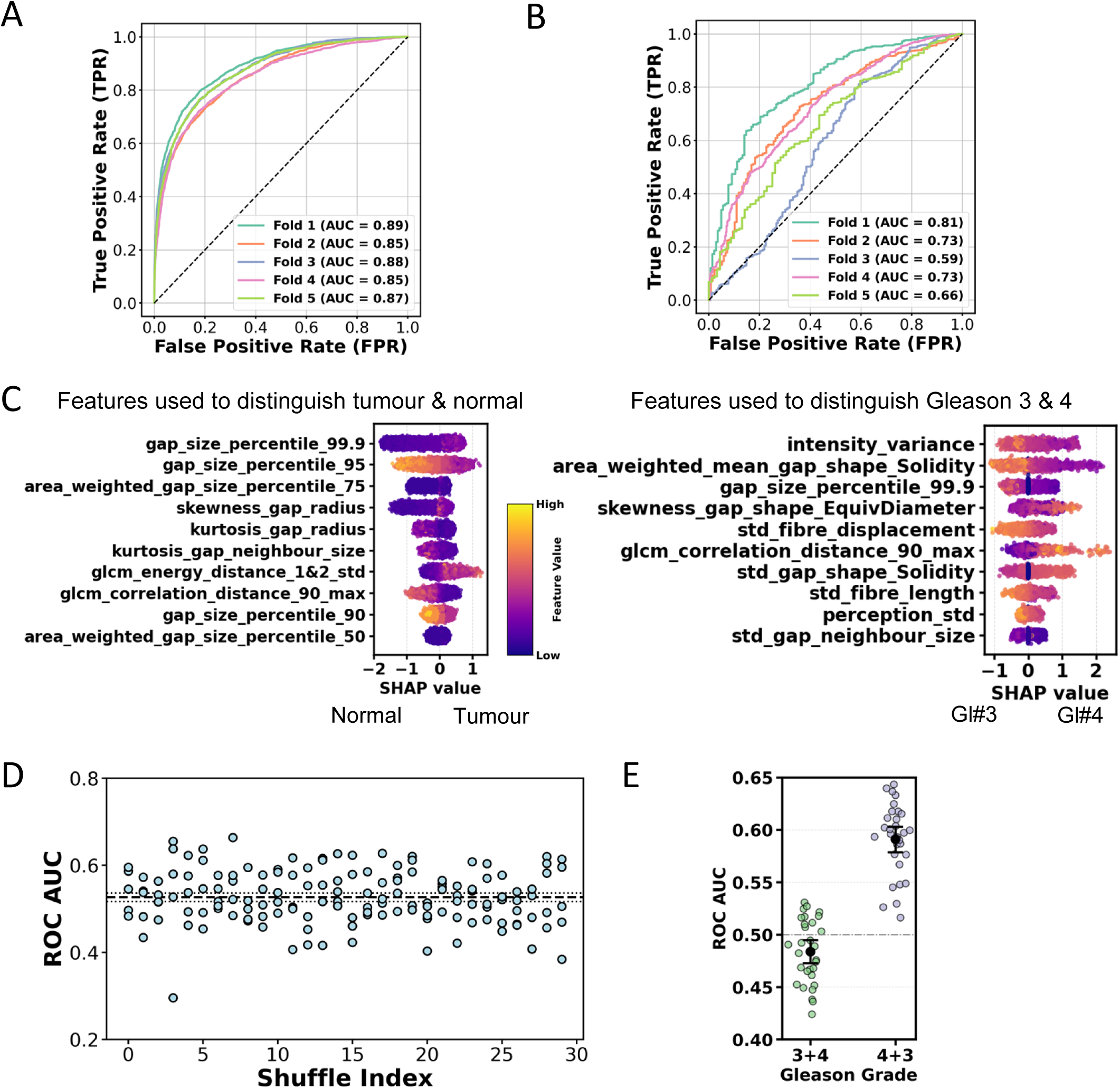
Tile level performance from decision trees. A: Receiver Operating Characteristic (ROC) curves for five outer folds of nested cross validation (CV) for normal (<=10% tumour) versus tumour (>=90% tumour) tiles for gradient boosted trees. Mean ROC AUC (Area Under the Curve) is 0.87. B: ROC curves for five outer folds of nested CV for Gleason 3 tiles (either from Gleason 3+3 WSI or demarcated by pathologist) versus Gleason 4 tiles (either from Gleason 4+4 WSI or demarcated by pathologist) for gradient boosted trees. Mean ROC AUC is 0.70. C: Aggregated Shapley values across all outer folds for normal versus tumour (top) and Gleason 3 versus Gleason 4 (bottom). D: ROC AUCs across all five outer folds of 30 random shuffles of patient fold designation for tile level recurrence for gradient boosted trees. Dashed line gives mean ROC AUC of 0.53, dot- dash lines give 95% confidence intervals of the mean generated from bootstrapping the mean of each shuffle 10000 times. E: Mean ROC AUC across folds for each shuffle separated by Gleason grade. Black dots and error bars give means and 95% confidence intervals from bootstrapping. Performance on Gleason 4+3 recurrence is ∼0.1 higher than for Gleason 3+4.

We then used the same approach to predict whether ECM features could identify Gleason pattern 3 versus 4. To do this, we used all patients with Gleason 3+3 and Gleason 4+4 histology and a pathologist (SW) annotated regions of Gleason 3 or Gleason 4 on 3+4 and 4+3 patients. The gradient boosted tree again outperformed the random forest with a mean ROC-AUC of 0.7 (Figure 4B) compared to 0.65 for the random forest (Figure S4C). Again, this demonstrates that the features could be used to successfully identify Gleason 3 versus Gleason 4 regions albeit with a lower ROC-AUC. The lower ROC-AUC may reflect the well-established challenges of manual Gleason scoring (22, 25).

To understand which features had the biggest influence on the models’ decisions, we determined their Shapley (SHAP) values. Figure 4C shows aggregated Shapley output across all the folds. Each datapoint represents a tile with the SHAP value along the x-axis being positive if it made tumour or Gleason 4 more likely. The larger the value, the bigger the effect of this feature on the predicted Gleason grade. The colour of each point represents the value of that feature for that tile. Features defining gap size were the biggest influencers for normal versus tumour prediction. High values of 99.9% gap size percentile were predictive of tumour tissue (note magenta dots to the right and dark blue dots to the left Figure 4C). Conversely, high values of 95% gap size percentile were linked to normal tissue. Together, this indicates that an abundance of large gaps (95 percentile), but absence of extremely large gaps (99.9 percentile), is associated with normal tissue. The features that distinguished Gleason 3 versus 4 were more mixed and included both measures of gap shape and measures of difference in texture. Gap solidity measures the regularity of the gaps with lower regularity increasing the likelihood of Gleason 4. Higher variance in pixel intensity decreased the likelihood of Gleason 4 while greater correlation in pixel intensities 20 microns (or 90 pixels apart) was linked with increased likelihood of Gleason 4 (glcm_correlation_distance_90_max).

### Machine learning to identify tiles from patients with disease recurrence

Next, we sought to classify recurrence at the tile level. Classifying tumour versus normal or Gleason 3 versus 4 is significantly aided by the fact that a ground truth is available making training and classification simpler. However, a ground truth for recurrence does not exist at the tile level and so we let each tile inherit the class of the patient i.e. all tiles from a patient developing recurrence were labelled as recurrent. A further challenge is that we do not know for certain that any recurrence signal exists within a given biopsy specimen, nor what proportion of the biopsy will have that recurrent signal. This problem is exacerbated by the very wide range in number of tiles representing each patient (Figure S2D). Hence, to further increase robustness, subject to stratification constraints, patients were randomised to outer folds. We did this 30 times resulting in different patients and tiles in each outer fold (Table S1). Nested cross validation is thus carried out 30 times i.e. with 30 shuffles, enabling us to report 150 ROC-AUC values (five outer folds at the tile and patient level 30 times) and increasing confidence in results (Figure S4A). Shuffles to predict recurrence are shown in Figure 4D with a mean ROC-AUC of 0.53 and confidence intervals of 0.52-0.54. On average, gradient boosted trees outperformed random forest models in predicting recurrence, although performance was similar between the two methods (Figures S4D). Interestingly, Figure 4E shows mean ROC-AUC to predict recurrence split by Gleason grade group; performance is significantly better with Gleason 4+3 tiles versus Gleason 3+4. These data indicate that collagen organisation may have a bigger impact on the probability of recurrence in Gleason 4+3 disease.

### Machine learning models to predict recurrence at the patient level

So far, we have made predictions of recurrence for each individual tile, where each tile inherited the recurrence status of the patient. Each patient’s prostate biopsy image is composed of multiple tiles. We therefore combined the tile level probabilities into three patient level statistics namely mean, standard deviation and maximum of the tile probabilities associated to that patient. These patient level statistics were put into a logistic regression model to produce a patient level prediction of recurrence. As before, we ran nested cross validation with five outer and five inner folds and randomised the outer fold each patient was placed in, subject to stratification constraints, 30 times. The patients associated to each outer and inner fold in each shuffle are identical to those used at the tile level resulting in no bleed through of information. Logistic regression hyperparameters were tuned on the inner folds, as before, as well as the probability threshold to determine predicted class, the minimum number of tiles required to make a given patient prediction and the choice of patient level statistics within the model. Hyperparameters were tuned using a constrained optimisation procedure that prioritised recurrence while maintaining acceptable performance for non-recurrence (see Methods).

The performance and split per Gleason grade group of the best performing models is shown in Figures 5A-B. F1-score is a measure of performance that combines precision and recall by calculating the harmonic mean of these two parameters. The F1-score for recurrence averaged 0.60 (95% CI 0.59-0.61) and ROC-AUC average 0.59 (95% CI 0.57-0.6). Performance once again increased for patients with Gleason 4+3 compared to 3+4 with ROC-AUC for 3+4 being 0.55, increasing to 0.64 for 4+3. During training on inner folds, we trained lower and upper bound hyperparameters to maximise performance for patients with the lowest or highest predicted recurrence probabilities. This enabled us to compare the accuracy of predictions with high, intermediate, and low probabilities of recurrence. Figure S5A shows that model performance is concentrated in the tails of probability distribution, with accuracy in the tails (most confident non-recurrent or recurrent predictions) being close to 0.7 and the accuracy of the remaining patients being 0.5. These higher confidence predictions account for around 25% of all patients.

**Figure 5:**
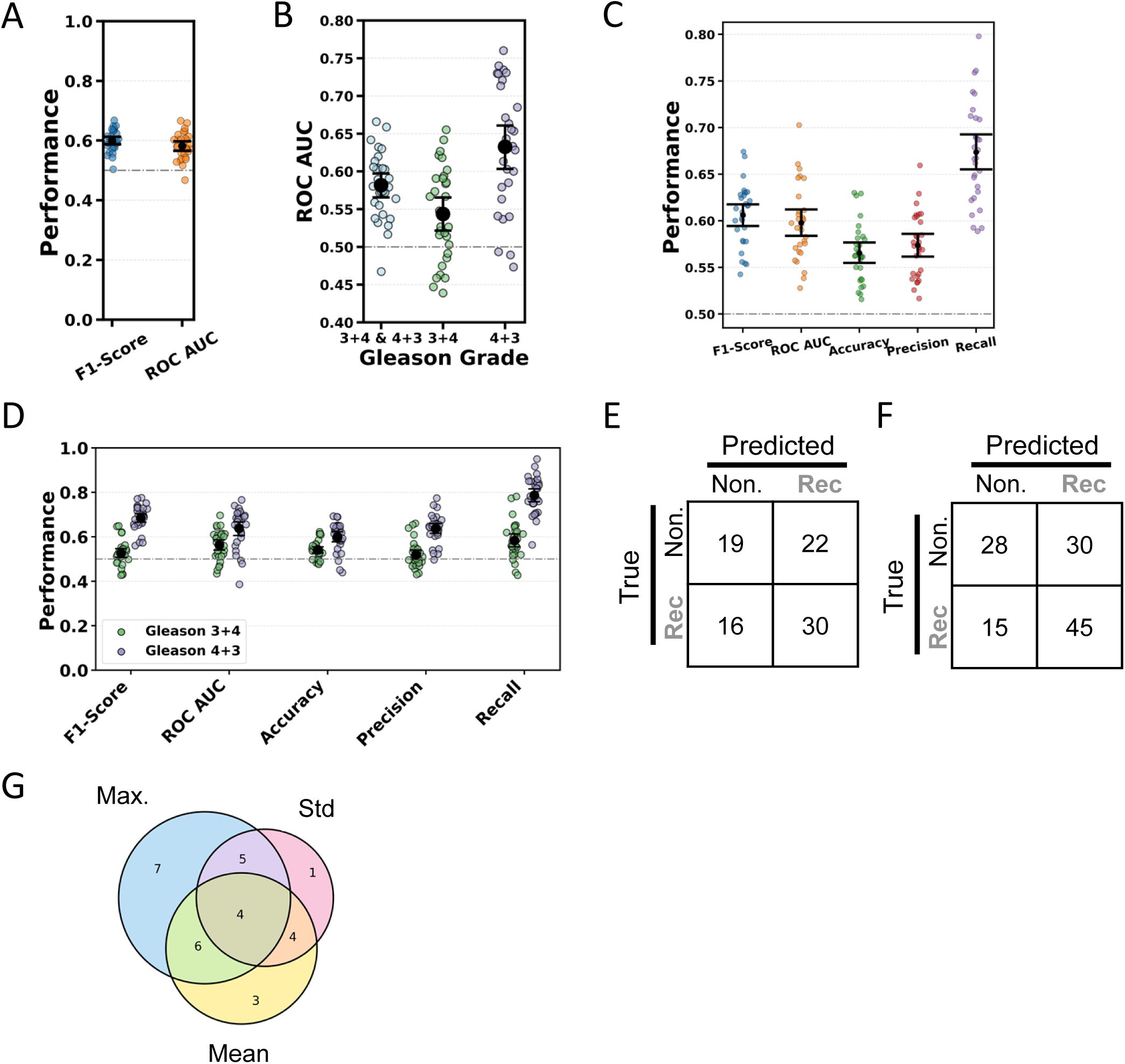
Patient level performance from logistic regression. A: Generalisation patient level performance metrics. Each dot is the mean performance across five outer folds of nested cross validation for a single shuffle. Error bars are 95% CI via bootstrapping across mean performance over all shuffles. Final probabilistic features used in logistic models are selected on inner loops leading to predicted generalisation performance of whole process. B: Splitting performance by Gleason grade shows the models’ performance is better than random in both cases but significantly better for Gleason 4+3 patients. C: Performance metrics having selected final models on outer loops for interrogation. A small amount of bias exists in prediction leading to slightly inflated values compared to 5A. All metrics are above 0.5 with recall being very strong i.e. the model is very good at identifying recurrent cases. D: Splitting performance by Gleason grade for models selected on outer loops. The model is better at identifying recurrent patients for Gleason 4+3 . E: Confusion matrix for sum over all five outer folds for mid performing shuffle. F: Confusion matrix for sum of all five outer folds for best performing shuffle. G: Venn diagram of patient level probability features used in final selected logistic model across shuffles.

Having initially demonstrated the generalisable performance of the full approach by considering patient level statistics selection on the inner folds, we next interrogated model performance by considering patient level statistics selection on the outer folds. This leads to a marginally over-optimistic prediction caused by using the outer folds for partial model selection. However, it has the advantage that a single model is selected on the outer folds leading to simpler model interrogation, with the single best model reported for each shuffle. Figures 5C and 5D show a range of performance metrics for the final selected models. Recall is particularly high – approaching 0.8 for Gleason 4+3, indicating that the model is very good at identifying recurrent cases for Gleason 4+3. Precision is holding down the F1-score suggesting some false positives for non-recurrence. Once again, performance is significantly higher for Gleason 4+3 patients than 3+4. Confusion matrices combined over all five outer folds for the mid performing shuffle and best performing shuffle are shown in Figures 5E and F, respectively.

For each patient we then calculated the proportion of the 30 shuffles that predicted that patient to be recurrent and related this to the accuracy of the prediction (Figure S5B). The majority of patients where greater than 90% of shuffles are deemed recurrent are recurrent, and all but one patients where less than or equal to 20% of shuffles (6 shuffles) are deemed recurrent are non-recurrent. This is presented in the confusion matrix of Figure S5C and demonstrates that when models consistently predict a patient to be recurrent or non-recurrent across shuffles, they are generally correct. This is consistent with the analysis in Figure S5A showing that the performance of the model is in the tails of the probability distribution.

We compared the performance of our models with the tumour cell intrinsic markers of PTEN status, maximum Ki67 positivity, mean Ki67 positivity, and Geminin expression, which are tumour suppressor genes or proliferation markers known to predict recurrence after prostate radiotherapy (24). These markers were available for a subset of patients in our cohort. We ran a cross validation on logistic regression providing ROC-AUC performance. These analyses yielded ROC-AUC values ranging from 0.53 to 0.59 (Figure S5D), indicating that our model is at least comparable to these validated markers. Unfortunately, because these markers only appeared on a subset of our data, we were not able to combine our ECM predictive approach with that of these markers.

Investigation of the metrics being used for patient level prediction by the 30 models revealed that 22/30 models used the maximum tile probability, 17/30 used the mean, and only 14/30 used the standard deviation of tile probability (Figure 5G). Figure S5E also highlights the greater ability of mean and maximum tile probabilities to distinguish between recurrent and non-recurrent patients. These analyses suggest that recurrence could be driven in part by a relatively small number of tiles with a clear recurrent signal (maximum) that also serve to elevate the mean.

### Explainability of patient level and tile level decisions relating to recurrence

Since no ground truth of recurrent signal exists, we next interrogated which spatial patterns drove the predictive models’ decisions, to shed light on ECM structures that associate with recurrence. We leveraged insights from having run nested cross validation 30 times where if specific tiles are repeatedly labelled as recurrent, regardless of shuffle, then we can be more confident that they incorporate recurrent signal. Figure 6Ai plots the mean probability of recurrence for each tile against the standard deviation of the probability across the 30 shuffles. We then sub-divided this plot with a grid and calculated the ROC-AUC using all tiles with a higher probability and lower standard deviation of recurrence than the grid point. Alongside this, we plotted the proportion of tiles, with a higher probability and lower standard deviation of recurrence than the grid point, that were recurrent (Figure 6Aii). This enabled visualisation of how ROC-AUC changes as the mean tile probability increased and the standard deviation of probability decreased. Specifically, we were able to identify tiles consistently linked to recurrence (yellow dots and red box Figure 6Aii, see also Figure S6A).

**Figure 6:**
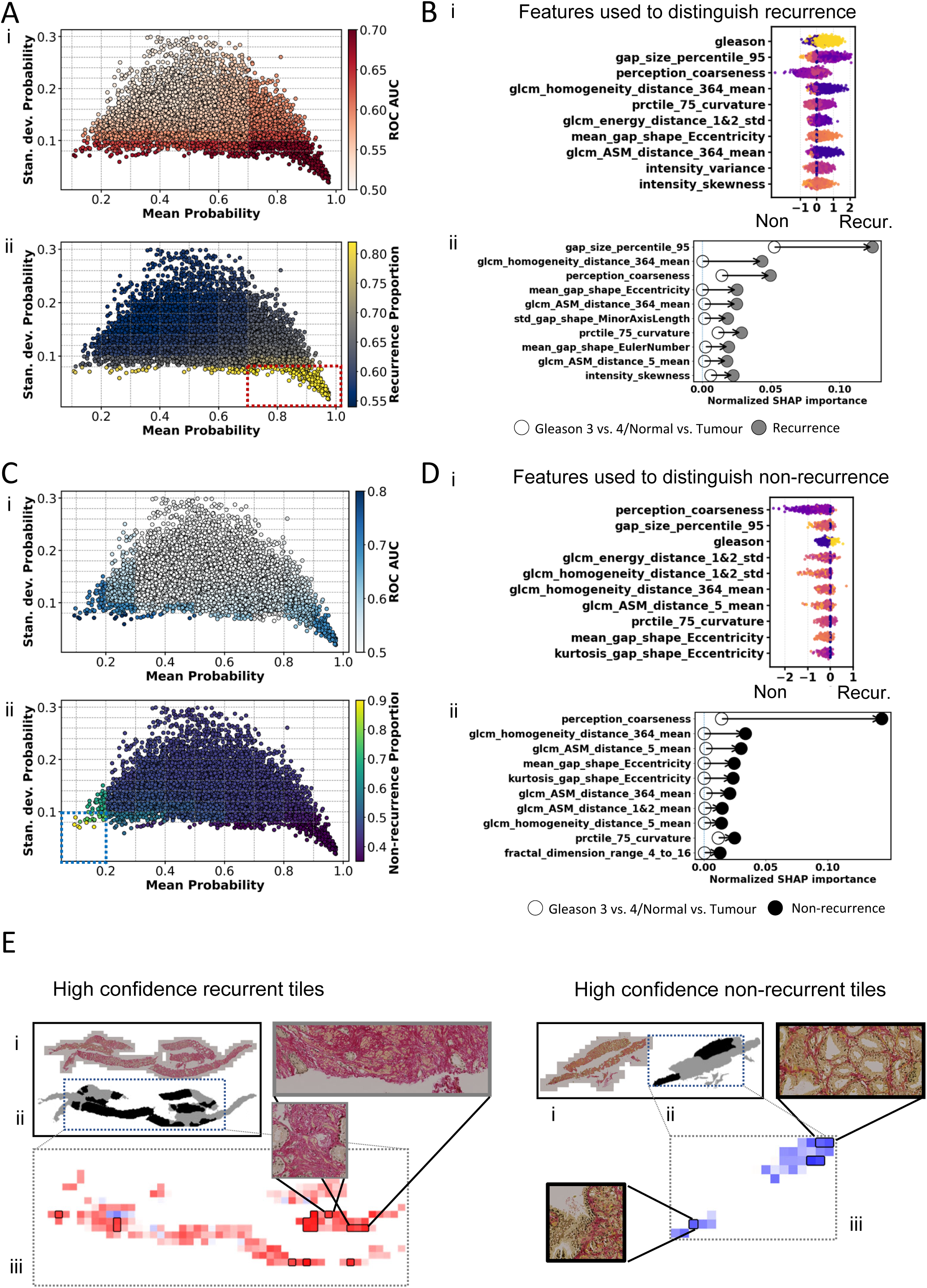
Explainability of tile level and patient level decisions: A: (i) Heatmap represents ROC AUC value obtained if considering only standard deviation less than and mean more than grid values. (ii) Heatmap represents proportion of recurrent tiles if considering only standard deviation less than and mean more than grid values. The highly recurrent region corresponds to the region with highest ROC AUC where std. dev <=0.08 and mean prob >=0.7. B: (i) Shapley values for region identified as highly recurrent, taken form all folds and shuffles and (ii) greatest normalised difference in Shapley values between recurrent classification and tumour/normal and Gleason3/4 classifications. The plot reveals those recurrent features independent of tumour or Gleason grade. C: (i-ii) Plots of mean tile level probability and standard deviation of tile probability over all 30 shuffles for all tiles. (i): Heatmap represents ROC AUC value obtained if considering only standard deviation less than and mean less than grid values. (ii) Heatmap represents proportion of non-recurrent tiles if considering only standard deviation less than and mean less than grid values. The highly non-recurrent region corresponds to std. dev <=0.1 and mean prob <=0.2. D: (i) Shapley values for region identified as highly non-recurrent, taken from all folds and shuffles. (ii) Greatest normalised difference in Shapley values between non-recurrent classification and tumour/normal and Gleason3/4 classifications. The plot reveals those non-recurrent features independent of tumour or Gleason grade. E: Exemplar WSI with highly recurrent/non-recurrent tiles highlighted. (i) The raw PSR+H WSI. (ii) The tissue and tumour mask (tumour in black, normal adjacent tissue in grey. (iii) Tile based heatmap showing mean probability of recurrence over all 30 shuffles and blocks highlighting highly recurrent/non-recurrent regions. Highly non-recurrent regions are more glandular, highly recurrent regions are more fibrotic.

Having identified tiles strongly linked to recurrence, we interrogated their Shapley values to determine which spatial features were dominating the decisions to strongly classify as recurrent (Figure 6Bi). Interestingly, Gleason grade occurs within these Shapley values. This relates to Figure 4E whereby the model had superior performance at predicting recurrence for Gleason 4+3 tiles. The models have ‘learnt’ that within Gleason 4+3 tiles, there is a specific ECM pattern for recurrent signal such that the model can ask in sequence “Is the patient Gleason 4+3?” followed by “does the tile also follow a (different) specific ECM pattern?”. If both questions are answered in the affirmative, the tile is most likely recurrent. The top Shapley values also included two ECM features describing gap size and shape, with low values of 95 percentile gap size linked to recurrence, and two describing image texture which point to areas of high-density ECM showing the strongest association with recurrence. To verify that these features are specifically identifying recurrence, rather than normal/tumour status or Gleason grade group, we derived the absolute difference in Shapley values between recurrence prediction and normal/tumour and Gleason grade predictions (Figure 6Bii). The resulting difference in Shapley values indicates there is substantial ECM signal that is specifically related to recurrence.

We next ran a similar grid-based analysis to define a region with low mean probability of recurrence, with a high proportion of non-recurrent tiles (80%) (yellow dots and blue box Figure 6Cii, see also Figure S6A). We then interrogated both the raw Shapley values and Shapley values that explain non-recurrence independent of Gleason grade and tumour (Figures 6Di & ii). ECM features that predicted non-recurrence were generally less extreme than those seen for recurrence. We observed that in this non-recurrent subset, Gleason grade was also a strong indicator with the reverse logic to above occurring such that the model asks in sequence “Is the patient Gleason 3+4?” followed by “does the tile follow this specific ECM pattern?” and if both are answered in the affirmative, the tile is most likely non-recurrent. We therefore identified a third region that was specifically Gleason 4+3 tiles with a strong non-recurrent signal with a standard deviation<=0.1 and mean probability <=0.6 (turquoise dots in Figure S6A). Reassuringly, many of the features identified by Shapley analysis for non-recurrence were also present here (Figure S6B, compare with Figure 6D). Imposing these highly (non) recurrent tile regions onto the WSI reveals pockets of recurrent or non-recurrent regions illustrating highly fibrotic (recurrent) or more glandular (non-recurrent) states (Figure 6E).

### Neighbourhood analysis of highly recurrent regions

Finally, in order to build up a fuller picture of regions driving recurrence, we analysed the spatial pattern of tiles that neighbour the highly recurrent tiles (Figure 7A). We plotted where these neighbours were, including frequency of neighbouring points (see colourmap Figure S7A) Most neighbouring points still had a high mean probability of recurrence but the standard deviation across shuffles increased, suggesting lower overall model certainty that the spatial pattern within that tile is strongly recurrent. Plots of the features that show greatest difference in Shapley values between the highly recurrent regions and neighbouring regions are shown in Figure S7A. Four features consistently emerge as important: Firstly, gap_size_percentile_95 i.e. how large are the 95^th^ percentile gaps; Secondly, mean_gap_shape_Eccentricity i.e. how rounded or elongated are the gaps; Thirdly, Perception_coarseness – how fine grained or uniformly smooth is the pixel texture and Fourthly, glcm_homogeneity_distance_364_mean – how similar does pixel texture look over a length scale of 80 microns (364 pixels). In combination, these suggest that highly recurrent regions had smaller more elongated gaps between fibres, with fine grained ECM texture and lower ECM homogeneity than less recurrent regions (Figure S7B). These four features also dominated in Figure 6B. Visualisation of exemplar highly recurrent tiles is shown in Figure 7B. They contain very small, elongated gaps between fibrotic tissue. In addition, the ECM exhibits a fine-grained texture that is not spatially consistent, such that regions separated by approximately 80 µm often differ in appearance. A correlation heatmap demonstrates that, apart from the GLCM and gap size metric, the above four features are poorly correlated and thus detect distinct aspects of the spatial patterns (Figure S7C).

**Figure 7:**
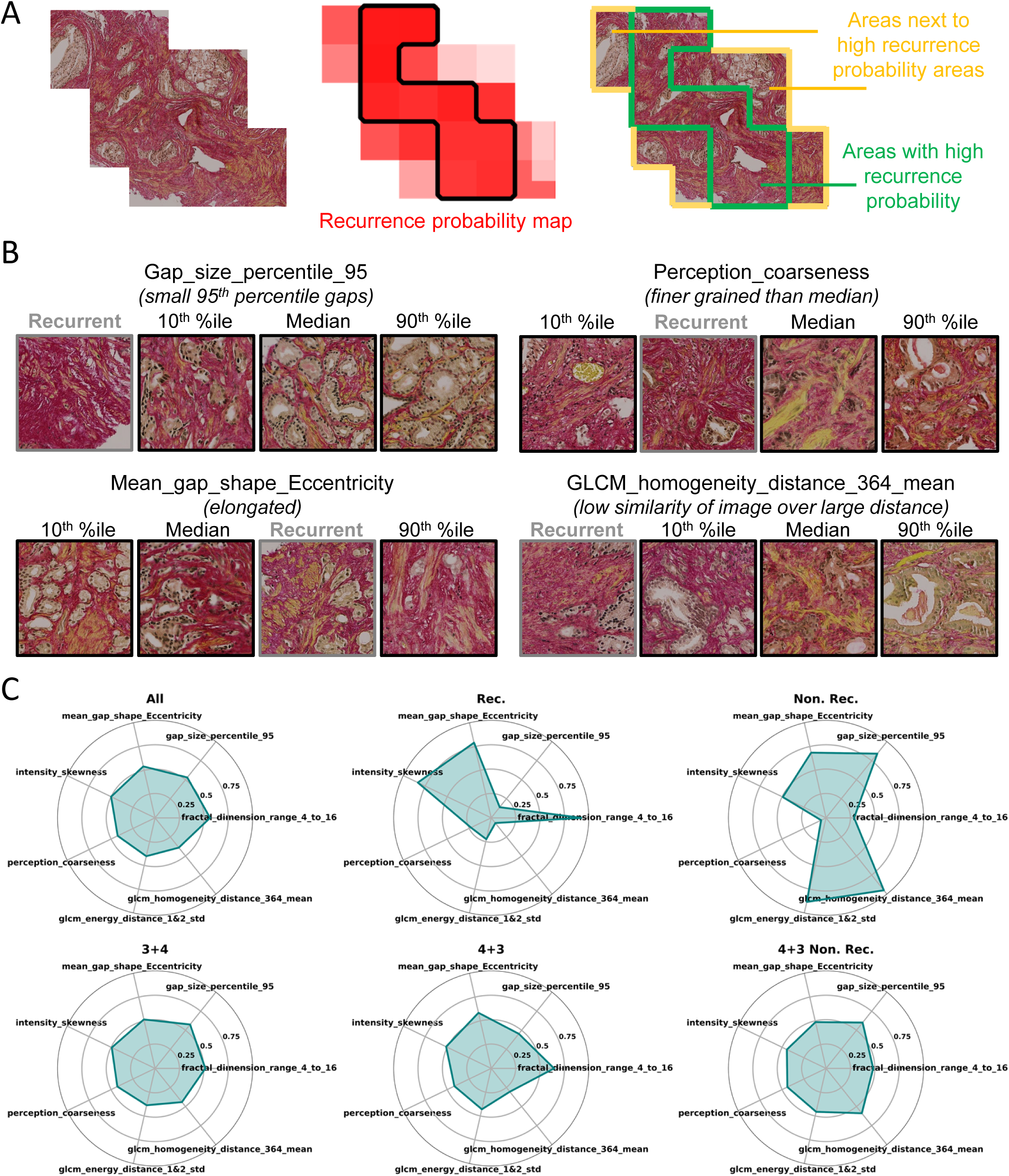
High recurrence neighbourhood analysis and outcome signatures: A: A section of recurrent WSI (left), recurrent map of mean probability of tile recurrence across 30 shuffles (centre) and the WSI with highly recurrent region marked in green and neighbours in yellow (right) for a single exemplar patient. B: Exemplar tiles for gap_size_percentile_95, mean_gap_shape_Eccentricity, Perception_coarseness and glcm_homogeneity_distance_364_mean for percentiles of all data demonstrating spread of pattern and median pattern for highly recurrent tiles. These four features are shown to be consistently the most diagnostic of highly recurrent regions. C: Radar plots for these four features plus three shown to be diagnostic of highly non-recurrent regions. All data, 3+4 and 4+3 all have similar signatures whilst highly recurrent and highly non-recurrent have distinctive signatures. Gleason 4+3 non-recurrence differs from Gleason 4+3 in gap size, fractal dimension and GLCM (Gray-Level Co-occurrence Matrix).

Lastly, we placed these features in radar plots (shown in Figure 7C) alongside three of the bigger hits for non-recurrence (Figures 6C&D). This enabled easy visualisation of the features that discriminate high and low probability of recurrence. Interestingly, these features did not show dramatic differences between Gleason 3+4 and Gleason 4+3, indicating that they provide distinct information relating to recurrence rather than Gleason grade and represent previously uncharacterised features of prostate tissue linked to recurrence following radiotherapy.

## Discussion

This study has generated a diverse set of quantitative features that comprehensively describe the ECM in localised prostate cancer and the surrounding normal prostate gland. We find substantial heterogeneity in ECM features within normal prostate tissue, within each Gleason grade group, and within individual prostate tumours, indicating a high overall complexity of ECM architecture. Our unbiased analysis identified ECM patterns that showed a moderate but robust predictive signal for identifying recurrence with high generalisability. Interestingly, most of the signal predicting recurrence arose in tumour areas, but ECM features derived from normal prostate tissue did show some signal which warrants further exploration. Our models were purely based on ECM features and recurrent and non-recurrent patient groups were balanced for Gleason grade group. As a result, the addition of other factors known to predict recurrence including Gleason, T stage, presenting PSA, proliferative index and oncogenic driver mutations such as PTEN loss (24, 26), may further improve predictive performance. We note that identifying an ECM signal of recurrence might be easier than non-recurrence as the former is linked to specific biological processes, whereas tissue states linked to non-recurrence are likely to be more diverse. Equally, challenges of identifying features predicting recurrence include that the relevant biological signal may not have been sampled at all in the core biopsies, and that the tumour phenotype driving recurrence may emerge after the diagnostic biopsy has been taken, for example during or after radiation.

We applied an unbiased discovery approach to the CHHiP cohort and acknowledge the need for wider validation of our findings in well-sized independent cohorts and across different stages of prostate cancer. Picrosirius red is an inexpensive and practical chemical stain however the ability to extract ECM signals from diagnostic H&E using generative artificial intelligence methods would further enhance the clinical utility of our models.

A challenge of diagnostic core biopsies, upon which treatment recommendations need to be made, is that defining the invasive tumour edge versus core can be difficult especially in multifocal disease. Intriguingly, the features we identify showing the most powerful prediction of recurrence arose in areas of low tumour cellularity which tend to arise at the tumour edge. The ECM features we identified that were specific to highly recurrence-associated regions including areas of dense aligned homogenous matrix with narrow elongated gaps show striking similarity with pathologist descriptions of “stromogenic” prostate cancer and preclinical studies that are known to associate with poor outcomes (8, 11).

Pathologists have recently incorporated the above stromogenic pattern into optimised definitions of adverse prostate pathology (27), but the subjective evaluation is a challenge for routine clinical implementation. Quantitative automated characterisation of stromal patterns, such as in this study, can overcome this challenge and additionally provide an opportunity to better understand matrix formation and tumour progression. Our methodology also represents a transferable tool of value to other biological problems. The practical advantages and relatively low cost of digital pathology AI algorithms means they are rapidly entering the clinic (28). Using such an algorithm, we demonstrate that the ECM architecture can provide clinically-relevant signals of recurrence in a common disease with challenging treatment decisions.

## Methods

### The CHHiP trial cohort

All patients provided written informed consent for the CHHiP trial and use of their tissue for research. The CHHiP trial, including the Trans-CHHiP (CRUK A12518) translational study, was approved in the UK by the London Multi-centre Research Ethics Committee (04/MRE02/10) and by the institutional research board of each participating international site. 2047 patients provided diagnostic biopsy tissue for translational research, that underwent central expert pathology review, including Gleason rescore (22).

### Definition of clinical cohort

In view of the fairly low recurrence rate in the CHHiP trial (20, 21), nested case: control methodology was used to define a cohort with 50% recurrence. Cases and controls were matched according to time to recurrence (i.e. each control was identified at the time of recurrence of a case) and Gleason grade group. As a result, the same patient could be both a control and then a case at a later timepoint. For recurrence classification, the small number of patients who fell in both groups were removed. Recurrence data was derived from a CHHiP data snapshot dated October 2020 where median follow-up was 9.2 years (interquartile range 8.2–11.0 years). Recurrence (BCR) was defined as biochemical failure incorporating the Phoenix definition (29), and/or clinical failure after radiation therapy. Non-recurrence was defined in patients with no evidence of BCR at the time of relevant matching to a recurrent patient. Patients were recruited to CHHIP from 71 centres. Following block collection, Gleason grade group was re-assigned centrally by a study specific uropathologist to reduce inter-observer variation and ensure grading was according to contemporary International Society of Urological Pathology (ISUP) guidelines (22). Tumour regions were manually defined on digital H&E and PSR images (aided by serial H&E and CK5/6 stained sections), images without tumour were excluded. Additional demarcation of Gleason 3 versus 4 for a subset of biopsy cases was performed in Qupath by a pathologist using both H&E and picrosirius red stained images.

### ECM staining

Interrogation of ECM organisation was enabled by picrosirius red and haematoxylin (PSR-H) staining of one 4μm section per patient using Abcam PicroSirius Red Kit (Connective Tissue Stain) (30). For this, the Weigerts haematoxylin stain is performed before the PSR step (following the manufacturer’s instructions). Slide immersion in acetic acid is performed for 1 minute exactly followed by a brief wash in ethanol prior to mounting. Subsequently, each slide was scanned at 20X using a Zeiss Axioscanner and tumour regions were defined manually on the resulting digital images.

### Tiling

WSI were gridded to enable extraction of 1000 pixel (220μm) square tiles from whole slide images (Figure 2a). The whole-slide image of PSR+H is read using aicsimageio Python library and converted into a Numpy ndarray. Tiling is performed by dividing the whole-slide Numpy ndarray to image patches, each with a dimension of 1000 x 1000 x 3. Features were extracted for tiles of sizes 440 microns squared (2000 pixels squared), 220 microns squared (1000 pixels squared) and 110 microns squared (500 pixels squared). Signal to noise ratio was determined to be highest for tiles of size 220 microns squared by observing increased effect-size.

### Colour deconvolution

Colour deconvolution was performed using the aicsimageio Python library to convert each image tile into 3 images, each with a dimension of 1000 x 1000 x 1. The stain map for colour deconvolution is determined in Fiji ImageJ by AW and ES, with values stain_color_map = {"psr": [0.174309, 0.8309804, 0.5282877], "nucleus1": [0.37535256, 0.61940926, 0.6852346], "nucleus2": [0.22552978, 0.6076955, 0.7614739]}. The “psr” channel after deconvolution, referred to as deconvolved PSR image, is used for downstream feature analysis.

### Tissue and tumour demarcation

To determine tissue boundaries, k-means clustering was applied to the RGB pixel values of 2.5x WSI. Non-tissue areas have a low variance in RGB channel. The majority of WSI neatly clustered into three groups – tissue and two background clusters. For WSI that failed this automated process, the correct number of clusters to extract tissue was assessed by visual inspection of reconstructed tissue signal. Following this, the tissue was binarized and the tissue boundary more smoothly determined using morphological opening and closing operations followed by MATLAB’s poly2mask function. Tissue masks and tumour annotations (Gleason 3, Gleason 4, and other tumour areas) were saved as grey-scale TIFF image files. Tissue and tumour annotations were combined by reading them as Numpy ndarray in Python. Tiling was performed concordantly with that used to tile the whole-slide PSR+H image. The tile-level deconvolved PSR image was filtered using the tile-level tissue mask image, through element-wise array multiplication, to create the masked-deconvolved PSR image. The proportion of tissue area within a tile and the proportion of tumour (Gleason 3, Gleason 4, other) within tissue area were calculated and linked with features at the tile level.

### Gap analysis

Circular gap analysis was conducted as in [23] to fit circles to the gaps in a binarized ECM mask of the greyscale deconvolved PSR image. Statistics including mean, standard deviation, skewness, kurtosis and percentile gap sizes are applied to these data and data weighted by circle area. Statistics regarding size of gaps neighbouring each other are also recorded. The discrete gap shape analysis was carried out by labelling gaps within the ECM mask as discrete connected components. Small regions of low intensity ECM can form bridges linking visually distinct gaps. To combat this, morphological closing was applied to generate a second more conservative labelling of gap regions. A distance transform was then used to split merged raw gaps according to the closest morphologically defined region. The method preserves small scale structure whilst generating more robust gap separation at the larger scale. These discrete gaps were then quantified both in raw form and weighted by area, using regionprops.

### Fibre extraction via CT-Fire

CT-Fire was used to extract discrete fibres (19). Tuning of optimal parameters was done for our data by a grid-search of key CT-Fire parameters and manual scoring of optimal fibre extractions by RPJ, XF, AW and ES. CT-Fire parameters were held at default except thresh_im2 at 106, s_xlinkbox at 2 and thresh_flen at 5.

### Fibre features

Discrete fibres were extracted from CT-Fire. Fibre straightness and shape metrics, by fitting alphaShapes to the fibres were derived. Statistics (mean, standard deviation, skewness, kurtosis and percentiles) were derived for curvature of all fibres. The angle of each fibre was calculated and circular statistics applied including angle standard deviation, skewness and kurtosis. Alignment of fibres a given distance away was calculated similarly to (31). Here, to increase efficiency we random sampled ECM points 1000 times and take both the median difference in angle between a point and its neighbours a given radius away and an equivalent method where points are weighted by ECM density. Fibre alignment was quantified 2.25, 4.5, 9, 23, 45, 68, 91 and 136 microns away. Mean, standard deviation, skewness and kurtosis of all alignment outputs were quantified. Branchpoints and endpoints were identified. Statistical tests comparing curvature at various regions were carried out. Fibre curvature and angle were extrapolated over the full ECM mask to test for directionality and uniformity of spatial distribution. High density matrix was calculated as ratios of fibres and ECM to tissue mask and fibres to ECM. Fractal dimension was calculated over length scales between 4.5, 18, 73,291 and 1164 microns. Length scales were selected appropriate to the tile size.

### Texture Features

Texture features were extracted from the masked-deconvolved PSR image. Three broad categories of texture features were considered: pixel intensity histogram, gray level co-occurrence matrix (GLCM) (32), and perception (33). The median, mean, variance, skewness and kurtosis of pixel intensity histogram were extracted using Numpy and Scipy.stats functions. The contrast, dissimilarity, homogeneity, energy, correlation and ASM of GLCM features were extracted using graycomatrix, graycoprops functions of the skimage.feature Python library. The coarseness and contrast of perception features were extracted using customised Python codes.

### Noise analysis and normalisation of features as a function of tissue proportion

Tiles with NaN values or complex values within features were removed. These all occurred for tissue proportion below 10%. The level of noise in feature values was assessed for varying tissue proportions by binning tiles into 100 bins from 1% tissue to 100% tissue. For each feature the coefficient of variation (ratio of standard deviation to absolute value of mean) was calculated and the median value across all features for each bin plotted. Tissue proportions below 0.2 were consistently shown to have high levels of noise, with an elbow in all plots being evident at this point. Feature values versus tissue proportion were fitted with a power law function and a power law function and linear regression fitted to data with at least 20% tissue. The MAE, MSE and R-squared values were optimal for power law fit. Hence a power function was fitted to all tiles for feature value versus tissue proportion. Feature values were then normalised to this power law via division.

### Feature removal process via correlation

Multiple features were shown to be highly correlated. Light touch feature removal was carried out by designating feature families and removing features that were highly correlated within these families. Feature families are shown in Figure 2D. Features with Pearson correlation greater than 0.999 were compared in terms of Cohen’s D effect-size for recurrence versus non-recurrence. Features with the lowest effect size were removed. Differences in effect-size for these features were typically 10^-3^ or lower. With this approach we removed 20 features for fibre and gap analysis. The greatest reduction in features occurred for GLCM where each feature was measured at multiple ranges and multiple angles. The redundancy and lack of rotation invariance of GLCM made us transform all features to being distributional. All features included standard deviation. All features except energy also included mean. Contrast, dissimilarity and correlation also included minimum and maximum at each length scale. Features at distance 1 and 2 pixels were combined. This reduced GLCM features from 216 to 136. The total number of features was therefore reduced from 478 to 378. We could have reduced them further but decided instead to allow feature selection to form part of the hyperparameter stage of ML model fitting (see below).

### UMAP

Hyperparameters were n_neighbors=5, min_dist=0.01, metric=’euclidean’ and random_state=42. For density representations the gaussian_kde function from scipy.stats was used.

### p-value and effect-size

For each feature, tiles were split into groups: tissue with less than 10% tumour (tumour adjacent) versus tissue with more than 90% tumour (tumour tiles); Gleason 3 (composed of tiles from Gleason 3+3 WSI and tiles with greater than 50% Gleason 3 in Gleason 3+4 and 4+3 as annotated by a trained pathologist) versus Gleason 4 (composed of tiles from Gleason 4+4 WSI and tiles with greater than 50% Gleason 4 in Gleason 3+4 and 4+3 as annotated by a trained pathologist) and recurrent tiles versus non-recurrent tiles. The t-test p-value and Cohen’s D effect-size were calculated with Benjamini & Hochberg multiple testing correction applied (multipletests from statsmodels).

### Data selection

For tumour versus normal we selected all tiles from all Gleason grades with tissue greater than 70% and compared tiles with <10% tumour versus those with >90% tumour. A total of 243 patients were included. For Gleason grade, tiles were selected with both tissue and tumour greater than or equal to 70%. We selected all Gleason 3+3 and Gleason 4+4 tiles and demarcated Gleason 3+4 and 4+3 tiles labelled as Gleason 3 or 4 based on dominant pattern. A total of 234 patients were included. For recurrence, we considered all Gleason 3+4 and 4+3 patients with tissue and tumour greater than or equal to 70% in selected tiles. A total of 228 patients were included.

### Nested cross validation

All models were trained using five-fold nested cross-validation (cv). Within each outer training set, five-fold inner cv was used to tune the model hyperparameters by selecting the combination that maximised the mean performance across the inner validation folds. The selected hyperparameters were then used to train a model on the complete outer training set, which was subsequently evaluated on the corresponding outer test fold. This process was repeated for each of the five outer folds, producing five independently trained models and five independent estimates of performance. The overall model performance was reported as the average across the outer test folds, providing an estimate of the model’s ability to generalise to unseen data. Because hyperparameter selection is performed entirely within the inner cross-validation, the outer test folds remain unseen until the final evaluation, avoiding optimistic bias in the performance estimate.

### Stratification

Stratification occurs at the patient level such that all tiles from a single patient will be in the same fold and the patients in each inner and outer fold are identical at both the tile and patient level. Patients are sorted by Gleason grade and recurrence. They are then sorted within each of these combinations in terms of number of tiles for each patient. For the outer folds, the fold numbers 1-5 are repeatedly randomly shuffled and concatenated to produce a list the same length as the total number of patients. The sorted list of patients are then assigned to an outer fold in the order of this concatenated random list. The same occurs for five lists of inner assignments. This method maintains splits between Gleason and recurrence grades across the folds to match the overall distribution. It also results in patients within each fold having as similar as possible distribution of number of tiles as the overall dataset.

### Patient level shuffles for nested cv

To further increase robustness to small numbers of patients with largely different numbers of tiles, the concatenated lists of random folds at the outer level are shuffled 30 times resulting in different patients and tiles in each fold for each shuffle at both the outer level and, as a result, the inner level but whilst maintaining the same distributions of Gleason grade, recurrence status and tiles per patient. Nested cross validation is thus carried out 30 times, resulting in us being able to report performance metrics for five outer folds at the tile and patient level 30 times, increasing our confidence in results. See Supplementary Table 1.

### Tile level Gradient boosted trees

The performance metric trained over was ROC-AUC with hyperparameter grids over the eligible range of hyperparameters of n_estimator =[100,200], max_depth=[3,5,7], learning_rate =[0.05,0.1], subsample=[0.7,0.85,1], colsample_bytree=[0.7,0.85,1], gamma = [0,0.5,1,5], scale_pos_weights = [1.8,2.3,2.8] and min_child_weights = [1,5,10] for tumour versus normal tissue, n_estimator =[100,200], max_depth=[3,5,7], learning_rate =[0.01,0.05,0.1], subsample=[0.7,0.85,1], colsample_bytree=[0.7,0.85,1], gamma = [0,1,5], scale_pos_weights = [0.1,0.2 1.0] and min_child_weights = [1,5,10] for Gleason 3 versus Gleason 4 and n_estimator =[100,200], max_depth=[3,5,7], learning_rate =[0.05,0.1], subsample=[0.7,1], colsample_bytree=[0.7,1], gamma = [0,1], scale_pos_weights = [1.0] and min_child_weights = [1,5,10] for recurrence of Gleason 3+4 or 4+3. Gradient boosted trees were implemented with xgboost’s XGBClassifier.

### Tile level random forests

Performance metric trained over was ROC-AUC with hyperparameter grids over the eligible range of hyperparameters of n_estimator = [25,50,100,200], max_depth = [3,10,20,100,None], min_samples_split = [2,5,10,20,50,100], min_samples_leaf = [1,2,5,10,20,50], max_feature = [0.05,0.1,0.25,0.5,0.75] and random_state = 8 for tumour versus normal tissue, n_estimator = [100,200], max_depth = [5,20,50], min_samples_split = [20,50,100,200], min_samples_leaf = [5,10,20,50], max_feature = [0.05,0.1,’sqrt’,’log2’] and random_state = 8 for Gleason 3 versus Gleason 4 and n_estimator = [100,200], max_depth = [5,20,50], min_samples_split = [50,100,200], min_samples_leaf = [10,20,50], max_feature = [0.1,’sqrt’,’log2’] and random_state = 8 for recurrence of Gleason 3+4 or 4+3. Random Forests were implemented with scikit-learn’s RandomForestClassifier.

### Tile level model performance

Due to the large number of features evaluated, as part of the hyperparameter tuning sci-kit learn’s RFECV feature reduction method was used to select only features that had some predictive signal. Model performance at the tile level was compared at the outer folds of nested cv and the selection of random forest or gradient boosted tree made on performance over all shuffles.

### Patient level agglomeration

Each tile for a given patient will have an associated probability of recurrence. For a given patient, the mean, maximum and sample standard deviation of probabilities of all tiles was calculated, with standard deviation set to 0 for patients with a single tile.

### Patient level logistic regression

Logistic regression models were trained for all possible combinations of three patient-level summary statistics (mean, maximum and standard deviation), giving seven feature sets in total. For each of these models, hyperparameters are tuned on the inner folds over the grid penalty = [’l1’, ’l2’, ’elasticnet’], C = [10**x for x in [-4, -3, -2, -1, 0, 1, 2, 3, 4]], solver = [’liblinear’, ’saga’], l1_ratio = [0.0, 0.25, 0.5, 0.75, 1.0] and max_iter = 1000 where ineligible combinations of hyperparameters are excluded. In addition, the minimum number of tiles required for inclusion of a patient in the patient-level analysis was treated as a hyperparameter. Patients with very few tiles may produce unstable patient-level summary statistics, potentially degrading predictive performance. The tile threshold was optimised over the range [0,2,5,10,15,25,30,35] within the inner cross-validation loop. The selected threshold was then applied unchanged to the corresponding outer test fold, and performance was reported only for patients whose tile count met or exceeded this threshold within each fold.

Hyperparameter selection prioritised accurate prediction of recurrence over non-recurrence. However, optimising solely for the recurrence F1-score resulted in poor performance for the non-recurrence class. Therefore, hyperparameters were selected using a constrained optimisation procedure that prioritised recurrence while maintaining acceptable performance for non-recurrence. For each hyperparameter combination, the mean recurrence and non-recurrence F1-scores were calculated across the five inner validation folds. Candidate hyperparameter combinations were first restricted to those for which the mean recurrence F1-score was at least as high as the mean non-recurrence F1-score. A minimum acceptable mean non-recurrence F1-score, p, was then imposed. The value of p was chosen as the largest value in {0.9,0.7,0.5,0.3,0.1} for which at least one hyperparameter combination satisfied the constraint, thereby enforcing the strictest feasible requirement on non-recurrence performance. Among the remaining candidate models, the hyperparameter combination with the highest mean recurrence F1-score was selected. Where multiple combinations achieved the same mean recurrence F1-score, the model with the highest mean ROC-AUC across the inner validation folds was chosen.

Model training was performed using nested cross-validation. For each outer fold, hyperparameters were tuned within the corresponding inner cross-validation loop. Within this inner loop, both hyperparameters and feature subsets (derived from combinations of mean, maximum, and standard deviation summary statistics) were optimised jointly based on the inner-fold performance criteria described above. The selected configuration of hyperparameters and features was then trained on the full outer training partition and evaluated on the held-out outer test fold. This procedure was repeated across five outer folds, producing five outer-fold performance estimates that were subsequently aggregated to provide an overall estimate of generalisation performance.

For model interpretation, a single feature representation was selected post hoc. Specifically, feature sets were compared based on their mean performance across the outer folds, and the feature set with the highest mean outer-fold performance was selected. This feature set was used for downstream model interrogation to ensure a consistent and interpretable representation of the final model structure. To assess stability of the modelling pipeline, the entire nested cross-validation procedure was repeated across 30 random shuffle splits of the data, yielding 30 repeated estimates of generalisation performance and 30 corresponding interpretable model configurations.

### Performance for low and high risk patients

Predicted probabilities from the trained models were analysed to quantify class enrichment in low, intermediate, and high-risk patients. Within each region, probability bins were constructed and class proportions were computed to characterise local class purity across the probability distribution. Analyses focused on the extreme regions of the distribution, with tail regions restricted to at most the upper and lower 25% of observations to ensure sufficient sample support while capturing distributional extremes. For each shuffle and nested outer fold, probabilities were partitioned into low-risk (≤L), high-risk (>U), and intermediate regions. Lower (L) and upper (U) probability cut-points were derived using inner cross-validation by evaluating candidate thresholds across probability bins based on class enrichment (proportion of true class membership within each bin). Robust thresholds were selected by maximising the median enrichment across inner folds, ensuring stability of tail definitions across resamples. The selected thresholds (L and U) were then applied to held-out outer-fold predictions. Predictive performance was summarised separately for the extreme regions (low + high combined) and the intermediate region, with accuracy used as the primary metric of overall classification performance within each region. In Figure S5A, each point represents the mean performance across outer folds for a single cross-validation shuffle. Individual outer folds may contribute no observations within a given region due to the application of L and U; therefore, point size reflects the number of outer folds within each shuffle that contained valid (non-empty) regions for evaluation. This provides an indicator of estimation completeness across the cross-validation pipeline.

### Patient shuffle recurrence analysis

For each patient, the held-out predicted class was recorded from each of the 30 repeated cross-validation shuffles. The proportion of shuffles in which the patient was predicted to have recurrence was then calculated. These proportions were displayed as frequency histograms for recurrent and non-recurrent patients (Figure S5B). For each histogram bin, the frequency represents the number of patients whose predicted recurrence proportion fell within that interval.

### Shapley Analysis

Shapley (SHAP) values were used to determine the features driving decision making at the tile level for normal versus tumour, Gleason 3 versus Gleason 4 and recurrence versus non-recurrence. SHAP values were computed independently within each cross-validation fold and mapped into a unified feature space defined by the union of all RFECV-selected features. For each fold, a full feature matrix was constructed over this global feature set. Features not selected in a given fold were assigned a SHAP contribution of zero for all samples in that fold, effectively encoding their absence as a null contribution within the model-specific explanation space. This allowed aggregation of SHAP values across folds while preserving a consistent feature ordering and enabling construction of a consensus feature importance and distributional SHAP summary. For the recurrent non-recurrent analysis over 30 shuffles, the same process was applied with each tile now represented 30 times, once per shuffle.

Shapley values for recurrence were compared to Shapley values for tumour versus normal and Gleason grade first via the aggregations described above. Mean absolute Shapley values were calculated for each feature providing a global measure of feature importance. These values were normalised within each classification task to obtain proportional feature contributions. To quantify recurrence-specificity, each feature’s normalised importance in the recurrence model was compared against the maximum corresponding importance observed in the tumour and Gleason models. A specificity score was defined as the difference between recurrence importance and the strongest competing classification task importance, such that positive values indicate features disproportionately associated with recurrence prediction. Features with positive specificity were ranked and visualised, highlighting features uniquely informative for recurrence.

### Identification of (non-) recurrent signal

Tile level recurrence probabilities differ over shuffles. The mean probability and standard deviation over shuffles was calculated for each tile. Tiles with high mean probability and low standard deviation of probability were deemed likely to have recurrent signatures. Using the mean probability for each tile, the ROC-AUC was calculated for all tiles with mean>=P_thresh and st. dev.<= std_thresh to determine regions with high performance at determining recurrence. Further regions of tiles for non-recurrence and Gleason 4+3 non-recurrence were also identified. The Shapley values for tiles in each of these subgroups were quantified as above. See Figure 6 and Figure S6.

### Recurrent neighbourhood analysis

All tiles that were 4-connected neighbours to highly recurrent tiles were identified. For subgroups of these highlighted in Figure S7, SHAP values were calculated as above. SHAP values were compared to the recurrent region for each neighbour subgroup. For each feature, mean SHAP magnitude was computed separately for the recurrence and neighbour subgroups, and the difference in these values was used to identify features with the greatest between-group difference.

### Software

Fibre and gap analyses were conducted in MATLAB R2019a, while k-means clustering for tissue demarcation was performed in MATLAB R2023b. All remaining analyses were implemented in Python 3.8.18. The following Python libraries and versions were used: *matplotlib* 3.7.3, *NumPy* 1.24.3, *pandas* 2.0.3, *scikit-learn* 1.2.2, *SciPy* 1.10.1, *seaborn* 0.13.2, *SHAP* 0.43.0, *statsmodels* 0.14.1, *UMAP-learn* 0.5.4, and *XGBoost* 2.0.0.

### Funding and Acknowledgements

This manuscript represents independent research supported by the National Institute for Health Research (NIHR) Biomedical Research Centre at The Royal Marsden NHS Foundation Trust and the Institute of Cancer Research, London. The views expressed are those of the author(s) and not necessarily those of the NIHR or the Department of Health and Social Care. The authors acknowledge funding from Cancer Research UK RadNet at The Institute of Cancer Research and The Royal Marsden Hospital and City of London RadNet. This project also received funding from a Crick i2i translational grant and Prostate Cancer Research. The CHHiP trial was supported by Cancer Research UK (C8262/A7253, C1491/A9895, C1491/A15955, SP2312/021), the Department of Health, the National Institute for Health Research (NIHR) Cancer Research Network, and NHS funding to the NIHR Biomedical Research Centre at the Royal Marsden NHS Foundation Trust and The Institute of Cancer Research, London. We thank the patients and all investigators and research support staff at the participating centres of CHHiP. Recognition goes to all the trials unit staff at the Bob Champion Unit and at ICR-CTSU who contributed to the central coordination of the study. We would also like to thank the CHHiP Trial Management Group, Trial Steering Group and Trans-CHHiP committee members.

A.W. is funded by a CRUK Clinician Scientist Fellowship (RCCCSF-Nov24/100003). E.S. is supported by the Francis Crick Institute, which receives its core funding from Cancer Research UK (CC2040), the UK Medical Research Council (CC2040), and the Wellcome Trust (CC2040) and the European Research Council (ERC Advanced Grant CAN_ORGANISE, Grant agreement number 101019366).

## Data Availability

All data produced in the present study are available upon reasonable request to the authors

---

**Supplementary figure 1:**
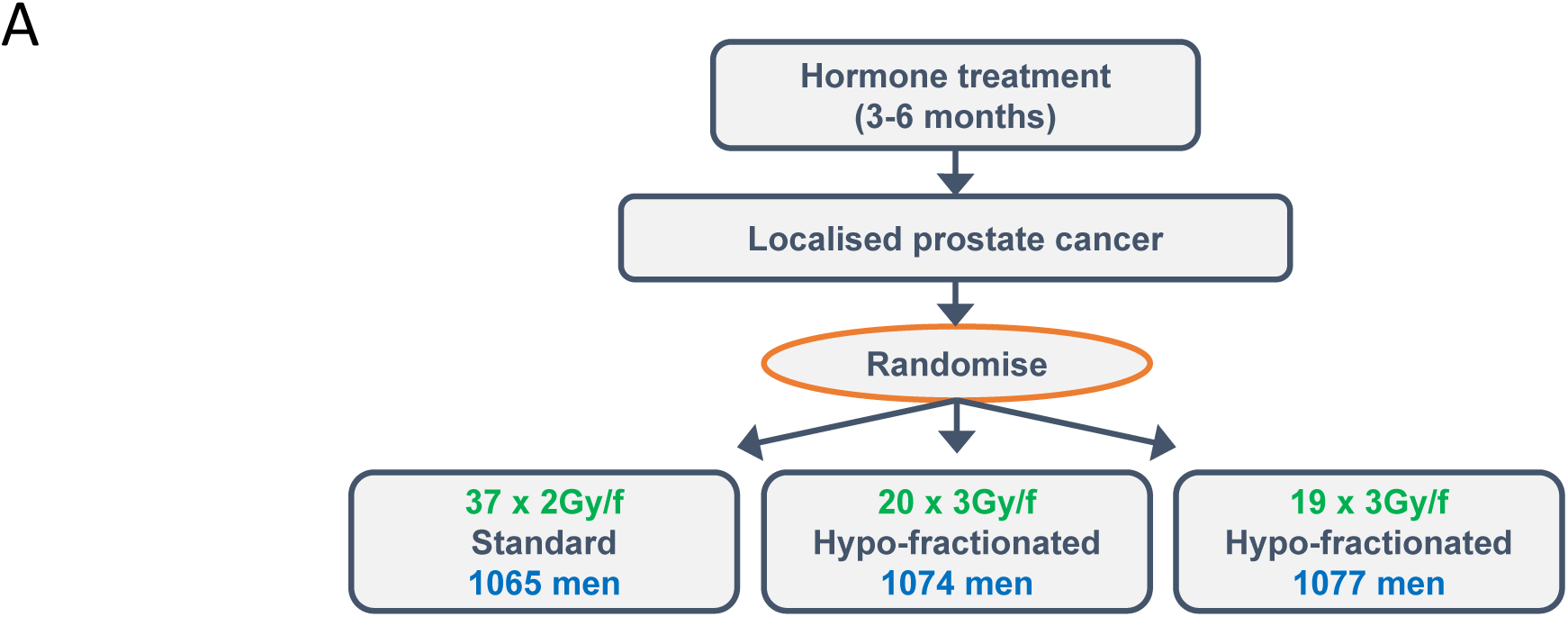
Schematics of clinical trial and machine learning pipelines. A: Full schematic of CHHiP trial.

**Supplementary Figure 2:**
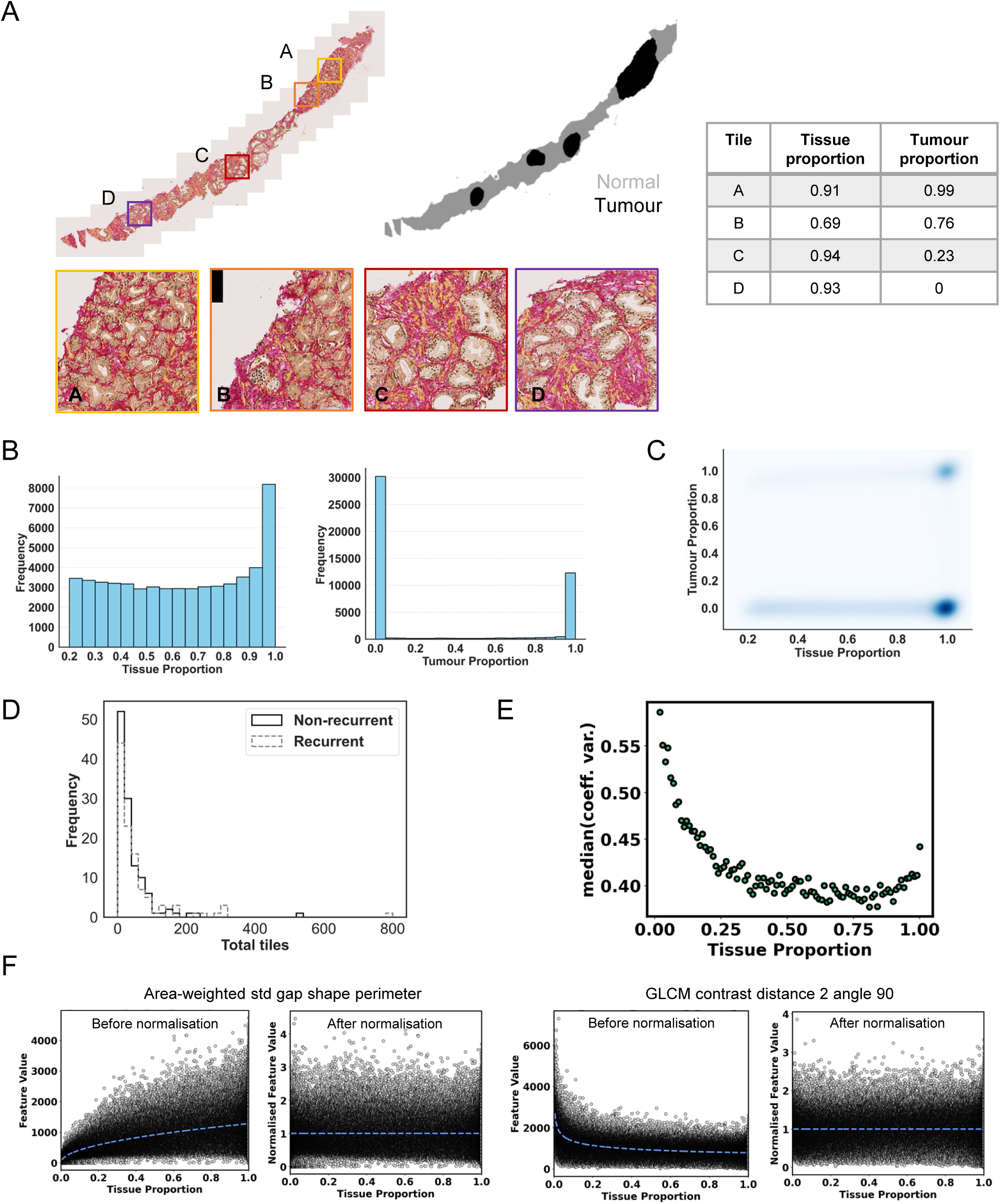
Distribution of data, pre-processing steps and feature generation. A: Illustration of tiling and tissue and tumour proportions. B: Frequency histogram of tissue proportion per tile for all tiles with at least 20% tissue and frequency histogram of tumour proportion per tile for all tiles with at least 70% tissue. C: Kernel density estimate of tissue and tumour proportions for all tiles with at least 20% tissue. Most tiles fall in two poles – high tissue proportion and high tumour proportion or high tissue proportion and low tumour proportion. D: Frequency histogram of number of tiles per patient for all Gleason 3+4 and Gleason 4+3 patients. E: Tiles are grouped into single percentiles of tissue proportion, the coefficient of variation calculated for each feature for tiles in that percentile and the median then plotted. There is a clear elbow at 25% tissue proportion showing that noise significantly decreases for tiles with at least 25% tissue. F: Features were normalised to tile tissue proportion by fitting a power law function. Left panel shows before normalisation and right panel after normalisation.

**Supplementary Figure 3:**
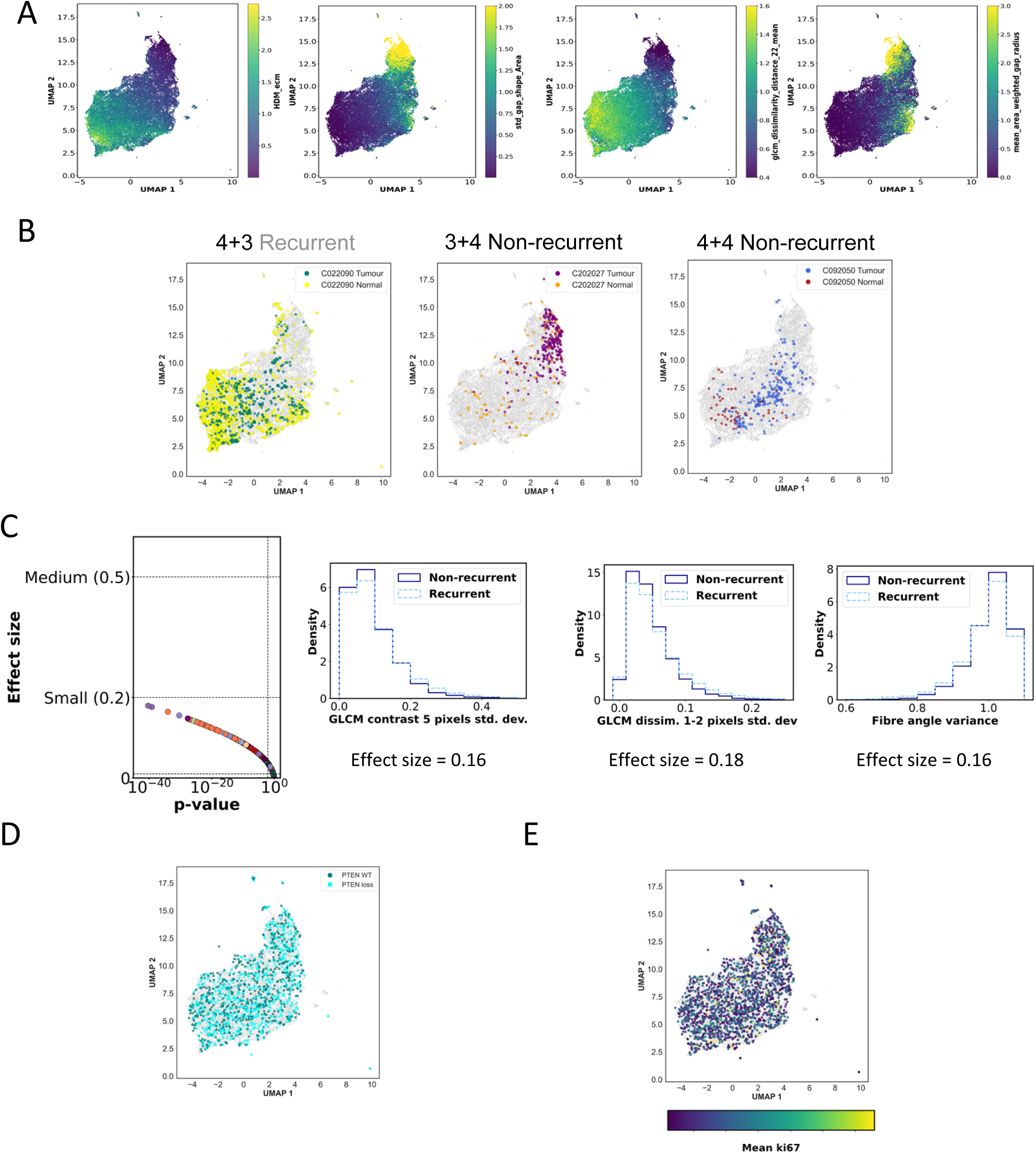
Tile level data exploration. A: Four spatial features overlaid onto UMAP space. High density regions occur predominantly in the bottom left. B: Exemplar plots show the distribution of normal and tumour tiles from three different patients. C: Plot of False Discovery Rate (FDR) corrected p-value versus effect-size for all features of non-recurrence versus recurrence and histograms of three features with largest effect-size for normal tissue tiles. D: UMAP dimensionality reduction with patient level PTEN wild-type (WT) (dark turquoise) versus PTEN loss (light turquoise) overlaid where available. E: UMAP dimensionality reduction with patient level mean ki67 levels overlaid where available.

**Supplementary Figure 4:**
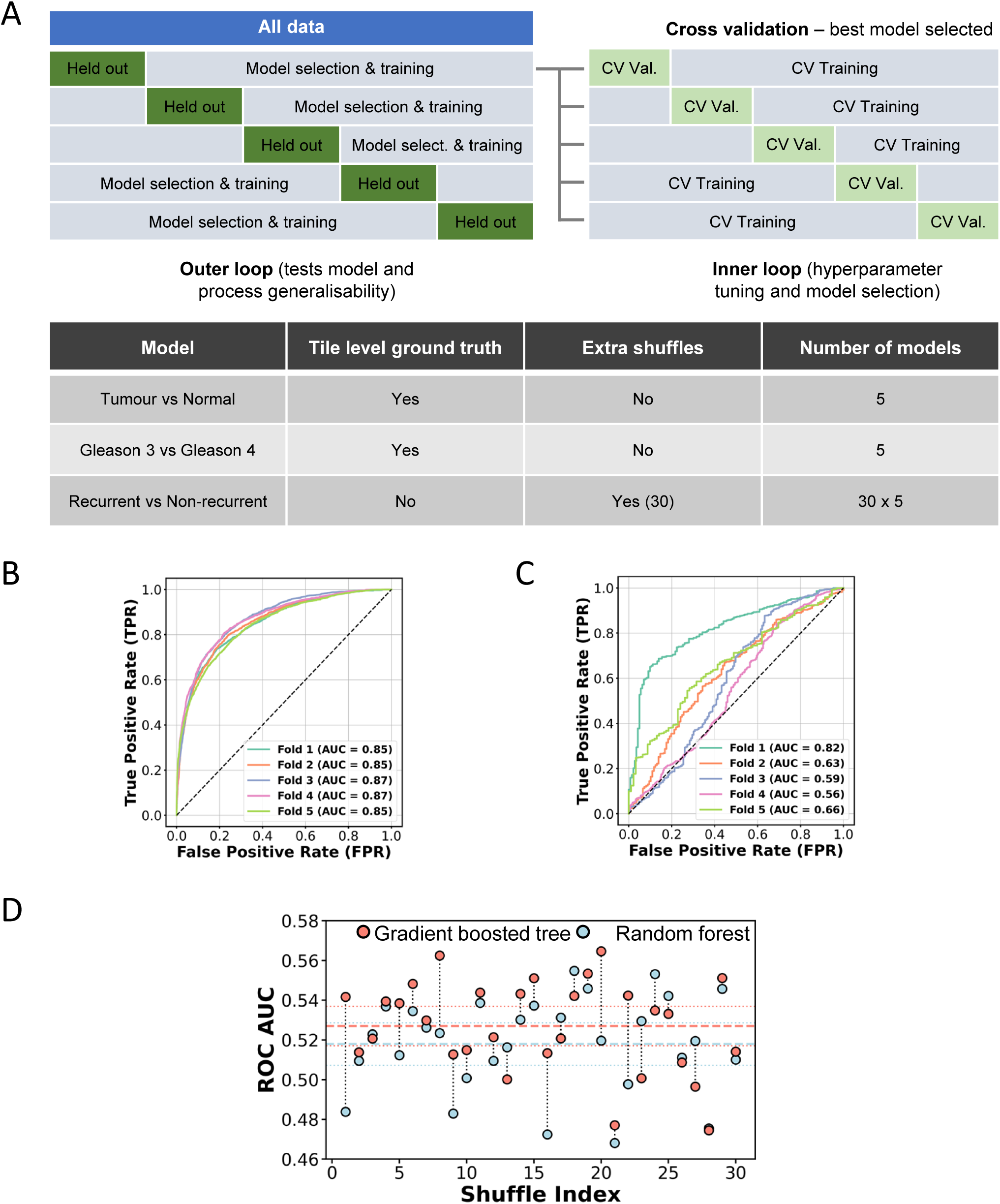
Tile level performance from decision trees. A: Schematic of nested cross validation (CV) and shuffling approach. B: Random forest ROC curves for five outer folds of nested CV for normal (<=10% tumour) versus tumour (>=90% tumour) tiles. Mean ROC AUC is 0.86. C: Random forest ROC curves for five outer folds of nested CV for Gleason 3 tiles (either from Gleason 3+3 whole slide image (WSI) or demarcated by pathologist) versus Gleason 4 tiles (either from Gleason 4+4 WSI or demarcated by pathologist). Mean ROC AUC is 0.65. D: Average ROC AUCs across the five outer folds of 30 random shuffles of patient fold designation for tile level recurrence for Gradient boosted tree (orange) and Random forest (blue). ROC-AUC: Receiver Operating Characteristic Area Under the Curve.

**Supplementary Figure 5:**
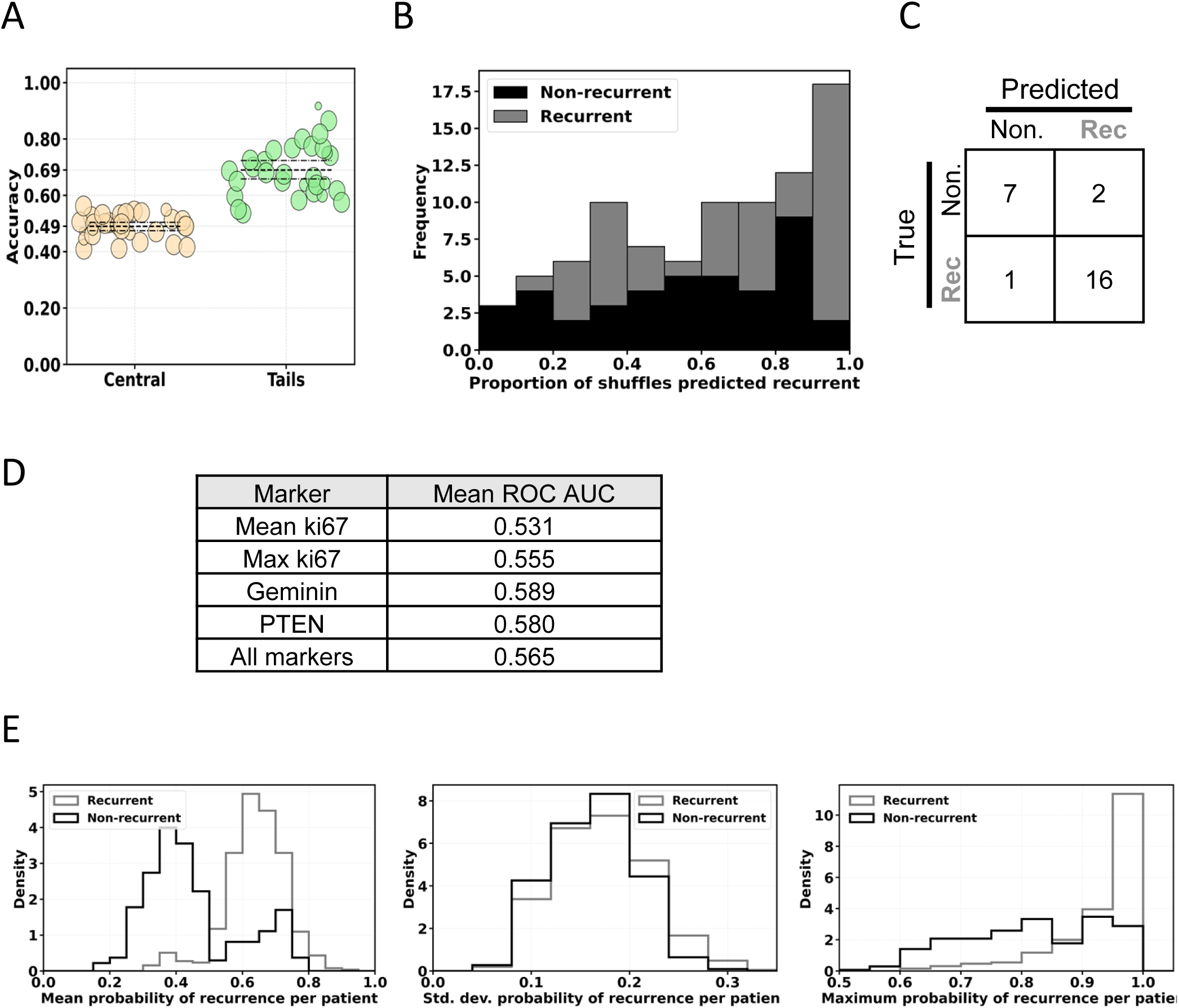
Patient level performance from logistic models. A: Performance of patients in tails of probability of recurrence have significantly higher performance than patients with recurrence probability close to middle of distribution. Here, upper and lower bounds to define tails were trained on the inner loops of nested cross validation and tested on the outer loops. Circle size reflects number of valid folds per shuffle (see Methods). B: Histogram showing frequency of patients where the proportion of shuffles in which the patient is classified as recurrent are between 0-0.1, 0.1-0.2 etc. Data is taken from outer loop fits as described in Figure 5C-G. For patients with >90% shuffles or <=20% predicting recurrence the results are dominated by recurrent and non-recurrent patients respectively. C: Confusion matrix just for patients with >90% shuffles or <=20% predicting recurrence demonstrating when the models consistently predict the same outcome they are generally correct. D: Logistic regression performance from 4 markers. Random forest used on case of all markers. E: Histograms showing differences in mean, standard deviation and maximum tile probabilities over all patients in the non-recurrent and recurrent groupings in SF5B&C. The biggest differences are in mean and maximum tile probabilities.

**Supplementary Figure 6:**
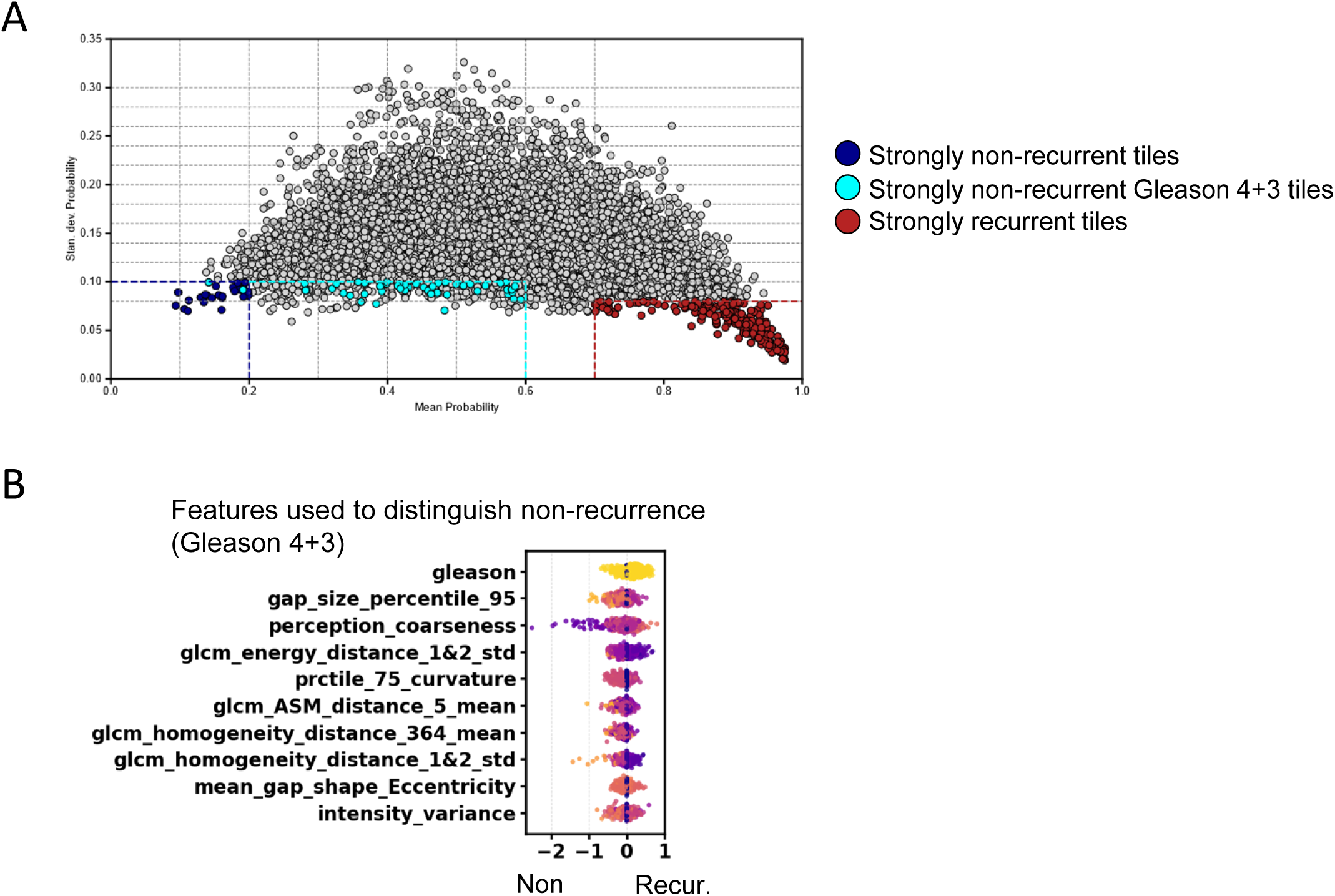
Explainability of tile level and patient level decisions: A: Plot of mean tile level probability and standard deviation of tile probability over all 30 shuffles for all tiles. Red region gives a subset of tiles where the model is highly confident of recurrence. Blue region gives a subset of tiles where the model is highly confident of non-recurrence. Cyan region gives a subset of Gleason 4+3 tiles where the model is confident of non-recurrence. B: Shapley values used to distinguish non-recurrence specifically in Gleason 4+3 regions.

**Supplementary Figure 7:**
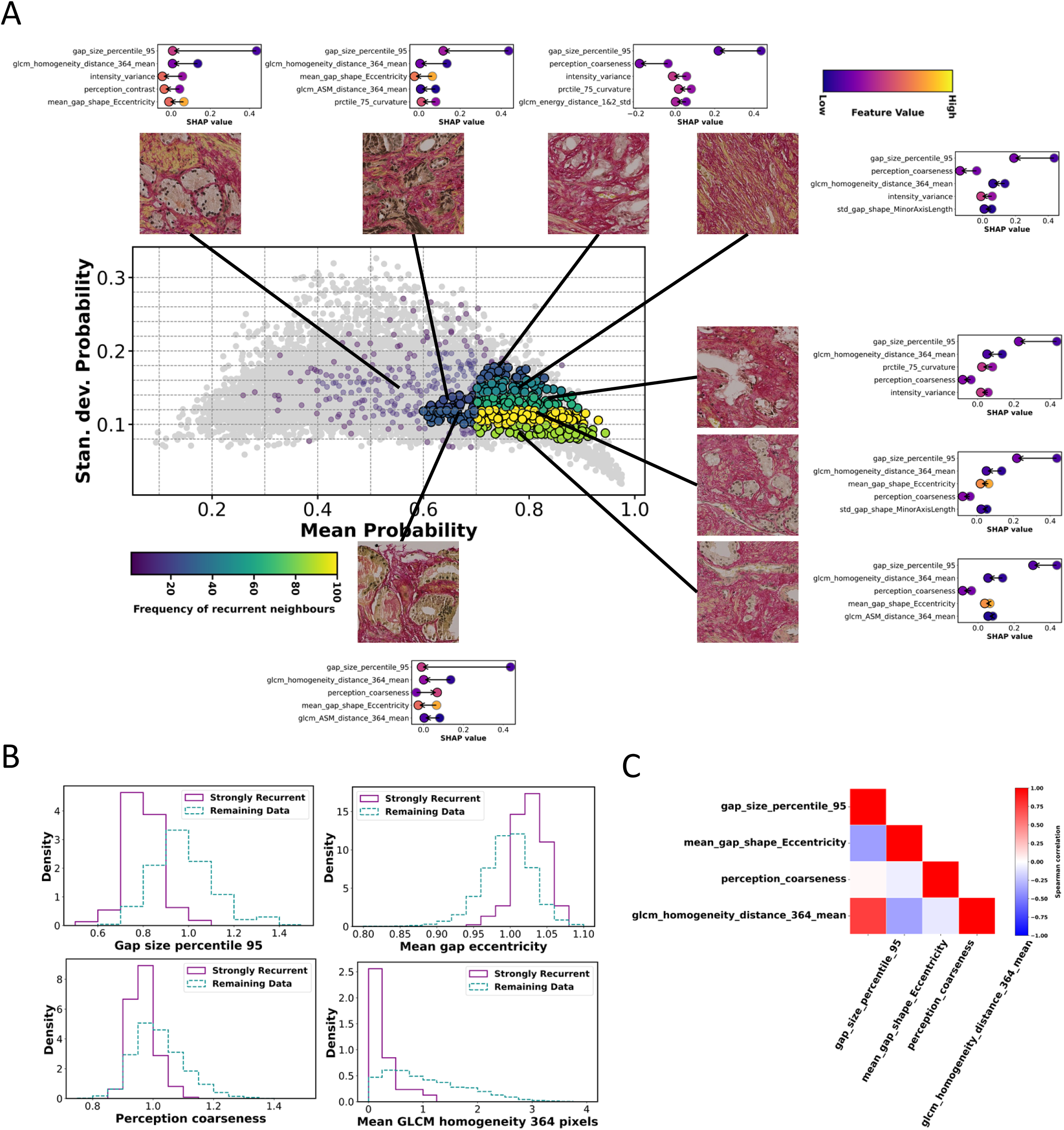
High recurrence neighbourhood analysis and outcome signatures: A: Scatter plot of mean probability of recurrence versus standard deviation of recurrence probability over all shuffles at tile level. Coloured points are neighbours to highly recurrent tiles with colourmap representing frequency of neighbours within each grid section. Exemplar tiles are shown in addition to mean difference in Shapley values between each region and highly recurrent tiles. The arrow for each feature goes in direction from highly recurrent region (grey disk) to neighbour regions (black disk). The colour within each disk shows the magnitude of that feature value, demonstrating how the feature shifts in different regions and drives a change in recurrence prediction. B: The four features consistently shown to have signal in (A) are gap_size_percentile_95, mean_gap_shape_Eccentricity, Perception_coarseness and glcm_homogeneity_distance_364_mean, all demonstrating the recurrent regions fall in an extreme for that feature. C: Spearman correlation shows that the four features are overall poorly correlated and pick up different aspects of spatial pattern.

**Supplementary Table 1:** Exemplar table demonstrating patient stratification across Gleason grade, recurrence status and then by number of tiles per patient. Each patient is randomly placed into one of five folds based on this ranking. For recurrence, the fold allocation is randomly shuffled 30 times for outer folds, generating 30 versions of five fold nested cross validation.

| Patient | Gleason | Recurrence | No. Tiles | Fold Shuffle 1 | Fold Shuffle 2 | Fold Shuffle 3 |
| --- | --- | --- | --- | --- | --- | --- |
| 13 | 3+4 | Recur | 700 | 1 | 2 | 5 |
| 17 | 3+4 | Recur | 480 | 4 | 3 | 4 |
| 3 | 3+4 | Recur | 370 | 5 | 5 | 2 |
| 22 | 3+4 | Recur | 279 | 2 | 1 | 1 |
| 25 | 3+4 | Recur | 130 | 3 | 4 | 3 |
| 18 | 3+4 | Recur | 80 | 3 | 5 | 1 |
| 20 | 3+4 | Recur | 37 | 5 | 3 | 3 |
| 23 | 3+4 | Recur | 20 | 1 | 2 | 5 |
| 10 | 3+4 | Recur | 4 | 2 | 1 | 2 |
| 19 | 3+4 | Non-Recur | 300 | 4 | 4 | 4 |
| 15 | 3+4 | Non-Recur | 270 | 5 | 1 | 5 |
| 11 | 3+4 | Non-Recur | 260 | 1 | 4 | 4 |
| 24 | 3+4 | Non-Recur | 200 | 3 | 3 | 1 |
| 16 | 3+4 | Non-Recur | 120 | 2 | 2 | 3 |
| 31 | 3+4 | Non-Recur | 76 | 4 | 5 | 2 |
| 27 | 3+4 | Non-Recur | 40 | 4 | 2 | 2 |
| 12 | 3+4 | Non-Recur | 2 | 5 | 1 | 4 |
| 29 | 4+3 | Recur | 250 | 1 | 5 | 3 |
| 4 | 4+3 | Recur | 120 | 2 | 4 | 1 |
| 21 | 4+3 | Recur | 90 | 3 | 3 | 5 |
| 28 | 4+3 | Recur | 77 | 2 | 3 | 4 |
| 14 | 4+3 | Recur | 50 | 4 | 5 | 1 |
| 32 | 4+3 | Recur | 30 | 1 | 4 | 2 |
| 5 | 4+3 | Recur | 10 | 3 | 1 | 3 |
| 26 | 4+3 | Recur | 5 | 5 | 2 | 5 |
| 6 | 4+3 | Non-Recur | 600 | 1 | 2 | 5 |
| 9 | 4+3 | Non-Recur | 589 | 4 | 3 | 4 |
| 1 | 4+3 | Non-Recur | 400 | 2 | 5 | 3 |
| 30 | 4+3 | Non-Recur | 180 | 5 | 1 | 2 |
| 2 | 4+3 | Non-Recur | 20 | 3 | 4 | 1 |
| 8 | 4+3 | Non-Recur | 5 | 1 | 3 | 2 |
| 7 | 4+3 | Non-Recur | 2 | 4 | 4 | 3 |

